# Asymptotic analysis of optimal vaccination policies

**DOI:** 10.1101/2022.06.02.22275908

**Authors:** Matthew J. Penn, Christl A. Donnelly

**Affiliations:** Department of Statistics, MPLS Division, University of Oxford, Oxford, United Kingdom; MRC Centre for Global Infectious Disease Analysis, Department of Infectious Disease Epidemiology, School of Public Health, Faculty of Medicine, Imperial College London, London, United Kingdom

## Abstract

Targeted vaccination policies can have a significant impact on the number of infections and deaths in an epidemic. However, optimising such policies is complicated and the resultant solution may be difficult to explain to policy-makers and to the public. The key novelty of this paper is a derivation of the leading order optimal vaccination policy under multi-group SIR (Susceptible-Infected-Recovered) dynamics in two different cases. Firstly, it considers the case of a small vulnerable subgroup in a population and shows that (in the asymptotic limit) it is optimal to vaccinate this group first, regardless of the properties of the other groups. Then, it considers the case of a small vaccine supply and transforms the optimal vaccination problem into a simple knapsack problem by linearising the final size equations. Both of these cases are then explored further through numerical examples which show that these solutions are also directly useful for realistic parameter values. Moreover, the findings of this paper give some general principles for optimal vaccination policies which will help policy-makers and the public to understand the reasoning behind optimal vaccination programs in more generic cases.

**Author summary:** The COVID-19 pandemic has illustrated the importance of vaccination programs in preventing infections and deaths from an epidemic. A common feature of vaccination programs across the world has been a prioritisation of different groups within each country’s population, particularly those who are more vulnerable to the disease. Finding the best priority order is crucial, but may be complicated and difficult to justify to policy-makers and the public. In this paper, we consider two extreme cases where the best prioritisation order can be mathematically derived. Firstly, we consider the case of a population with a very small, very vulnerable group and show that this group should always be vaccinated first. Then, we consider the case of a small supply of vaccines and derive an equation which gives the best prioritisation order. Understanding these extreme cases is important, as it highlights general principles of optimal policies which will be useful when understanding the solution in more complicated settings.

## 1 Introduction

The trajectory of an epidemic can be dramatically changed by the implementation of a vaccination program, as has been shown in the case of COVID-19 [1]. These vaccination programs are most effective when they target specific groups in a population [2], although the optimal targeting strategy is dependent on the properties of the disease and vaccine [3]. Thus, it is important to have robust methods to determine the optimal strategy whenever a new epidemic emerges.

Used widely across the literature (in papers such as [4], [5] and [6]), the multi-group SIR (Susceptible-Infected-Recovered) model provides a general framework with which to assess the effectiveness of different vaccination policies. It splits a population up into a number of inter-connected subgroups (such as age groups) and captures the different transmission dynamics between each group. This construction highlights the dual benefit of vaccination as it both directly protects the individuals that are vaccinated and indirectly protects unvaccinated individuals by reducing transmission [7].

Often, especially in the case of age-structured populations, there is a negative correlation between the infectiousness of a group and the vulnerability of its members to the disease [8]. This means that the optimal strategy may not be obvious and indeed, the seemingly intuitive solution may be significantly sub-optimal [9]. Moreover, the complicated nature of the optimisation scheme, which involves solving the adjoint equations derived via Pontryagin’s Maximum Principle (see [10], [11]) means that the optimal solution may be difficult to understand or qualitatively justify to policy-makers.

When attempting to understand a complicated problem such as finding the optimal vaccination policy, it is often helpful to look at cases with extreme parameter values via asymptotic analysis, which helps the problem to be analytically solvable (at least to leading order). This can help form general principles for optimal vaccination policies. These principles can then be used both to form heuristics for finding the true optimal policy in a more general setting and also to explain the resultant optimal solution, as it is often comprised of a mixture of policies resulting from these principles.

There have been a number of recent papers that have used asymptotic analysis to derive general principles. [12] discusses a model with high reproduction numbers and shows that in this case, it is often optimal to vaccinate the less infectious groups in a population. Moreover, [13], building on the work of [14], linearises the model equations and derives a simple knapsack problem, although the solution to this problem is only optimal when considering the short-term evolution of the epidemic. Other special cases are investigated in [15] (which looks at a population with disconnected subgroups) and [16] (which examines the critical vaccination fraction for a population with separable mixing).

Despite this body of work, two cases will be considered in this paper which both provide novel contributions to the literature. Firstly, the case of a population with a small vulnerable sub-group will be analysed, and it will be shown that, in the asymptotic limit, any vaccination policy is eventually outperformed by one where this group is vaccinated first. Of course, the concept that vaccinating vulnerable groups is important has been raised in many previous papers, such as [3] and [17], but the mathematically rigorous asymptotics presented here provide new evidence for the importance of this principle.

The second case to be discussed is that of a a small total vaccination supply. The key novel result that will be shown is that (to leading order) the optimal vaccination problem reduces to a linear knapsack problem which can be easily solved. This knapsack problem differs from the one in [13] because, by linearising the final size equations rather than the model ODEs (ordinary differential equations), the optimal solutions and predictions of their behaviour are valid for the full evolution of the epidemic, rather than just in the short-term. Again, the case of a small vaccine supply has been examined in many papers such as [18], [19] and [20], but these papers have simply analysed the optimisation problem in the standard way, without deriving the explicit leading order solution as is done in this paper.

These analytic results will then be further investigated through examples and, in particular, the small supply case will be used to show that it is not always optimal to vaccinate the most infectious group, even when all groups are equally vulnerable. The UK population’s age structure will be used to relate these results to a realistic example, and optimal small-supply vaccination policies will be approximated for diseases with different age-dependent case fatality ratios.

The paper is structured as follows. Firstly, the multi-group SIR model will be introduced. Then, analytic results will be presented in the case of a small vulnerable subgroup, which will be explored through numerical examples. Finally, analytic results related to a small vaccination supply will be presented and again, examples will be used to illustrate the findings.

## 2 Modelling

### 2.1 Disease transmission and vaccination model

The model used in this paper is identical to the model presented in [24] and this section is simply a summary of the modelling section in [24]. The population is divided into *n* subgroups and each subgroup *i* is further divided into six compartments:

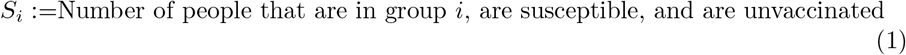

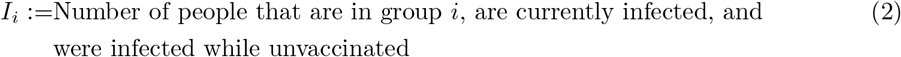

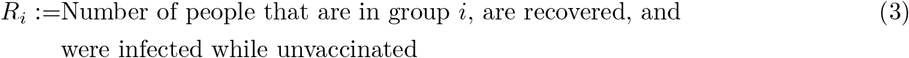

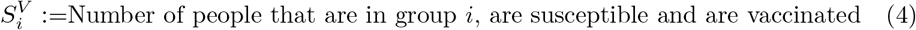

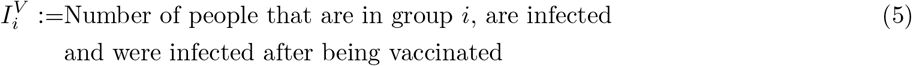

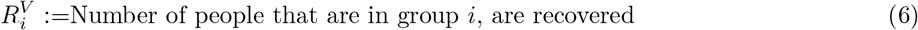

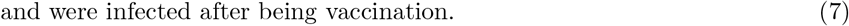

Using SIR principles, the model becomes

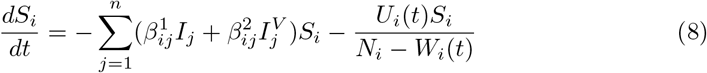

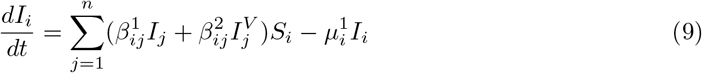

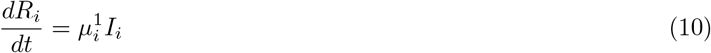

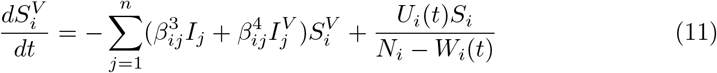

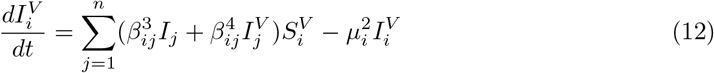

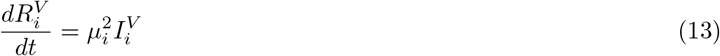

where

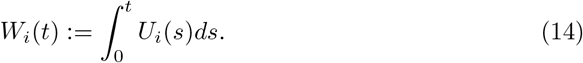

The 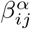 terms represent transmission from group *j* to group *i* and the 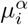 terms give the infectious period of the relevant individuals in group *i*.

Note that there is a slight difference between this model and the one commonly found in the literature (in [21], [22] and [23] among many others) which set the vaccination term equal to *S*_*i*_(*t*)*U*_*i*_(*t*) instead of 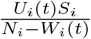. As discussed in [24], this corresponds to vaccines that are randomly distributed to the whole population, rather than the model in this paper which corresponds to vaccines that are randomly distributed only to the unvaccinated population. [24] provides justification for the use of the “unvaccinated-only model”, which is therefore the one that will be used in this paper. However, they are structurally very similar, and so it would be possible to apply the results in this paper to the more commonly found model.

It is worth noting that there is conservation of population in each group - summing the equations (8) to (13) and integrating gives

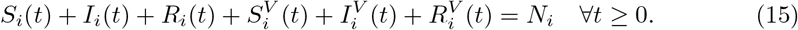

To deal with the (removable) singularity that can occur when *W*_*i*_ = *N*_*i*_, it is assumed that

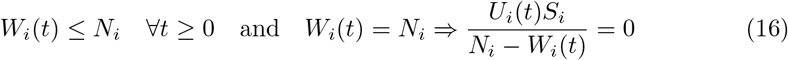

To capture the benefits of vaccination, there are additional constraints put on the 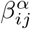 and 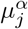 terms which are

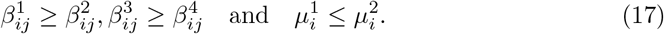

Again, details are given in [24].

### 2.2 Optimisation problem

The optimisation problem (which is again explained in more detail in [24]) is

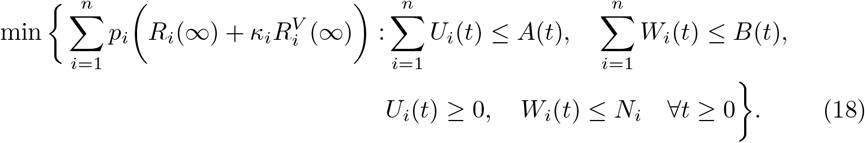

Here, *A*(*t*) represents the maximal vaccination rate and *B*(*t*) represents the maximal vaccine supply. It will be assumed throughout this paper that all “feasible” ***U*** are sufficiently smooth for all the quoted theorems to hold. In general, this does not significantly restrict ***U*** - for example, the results in [24] simply require that each *U*_*i*_(*t*) is bounded and Lebesgue integrable, while Theorems 1 and 2 require only that *U* has finite support. Moreover, it is assumed that *B*(*t*) is non-decreasing (as total supply should not decrease over time) and piecewise differentiable.

## 3 Results

### 3.1 A small, vulnerable subgroup

Consider the case where one of the groups in the population (which, without loss of generality, will be assumed to be group 1) is very small and vulnerable. That is, the size of the population *N*_1_ and the vulnerability *p*_1_ satisfy, for small *ϵ*

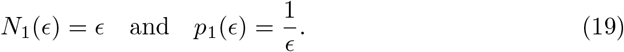

Note that, in the case that *p*_1_ is interpreted as a probability of a fatal infection (so remains bounded) this is equivalent to the case where

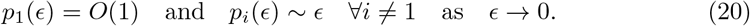

Note that by suitable rescaling, one can assume that 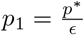 for any *p*^*^ > 0.

#### 3.1.1 Analytic results

The first result presented in this section shows that, in the limit of a group with small size and large vulnerability (with the total cost of the whole group being infected, *N*_1_*p*_1_, remaining constant) any fixed vaccination policy where the vulnerable group is not vaccinated first will eventually (that is, for sufficiently small *ϵ*) be outperformed by the equivalent policy where the vulnerable group is vaccinated first.

Group 1 will be given a population size *N*_1_ = *ϵ* and an infection cost 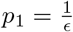. It will be assumed that the initial conditions in the group are proportional to *ϵ*, so that there exists some *σ* ∈ (0, 1] such that the initial susceptible population is *σϵ* and the initial infected population is (1 − *σ*)*ϵ*.

Before stating the full theorem, it is helpful to explain the various constraints and variables that will be introduced. Define, for each value of *ϵ* ≥ 0, ***U*** (*t*; *ϵ*) to be the “fixed” vaccination policy where group 1 is not vaccinated first. Of course, the vaccination policy cannot be completely fixed, as the size, *ϵ*, of group 1 is decreasing, and so it will simply be assumed that the vaccines given out to each group satisfies

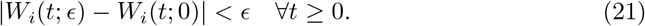

That is, any variation in the vaccination policy for non-zero *ϵ* is of size at most *ϵ*. Moreover, to reduce the lengths of the proofs, it will be assumed that ***U*** has uniformly bounded finite support - that is, there is some constant *t*_*U*_ such that for each

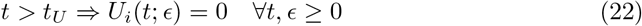

In order for group 1 to not be vaccinated first in the limit as *ϵ* → 0, there must be some time *τ* at which some fixed proportion *w* of the other groups have been vaccinated, while at least some fixed proportion (1 − *α*) of group 1 has not been vaccinated. That is,

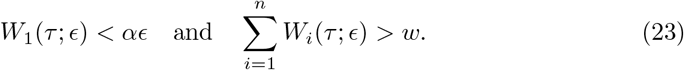

One can also define a vaccination policy ***Ũ***(*t*; *ϵ*) where group 1 is vaccinated first. This will be done by re-directing all vaccinations from the ***U*** (*t*; *ϵ*) policy to group 1 until it is fully vaccinated, and keeping the same vaccination policy after group 1 is fully vaccinated (ignoring any vaccines that ***Ũ***(*t*; *ϵ*) assigns to group 1 after this time).

To ensure convergence of the model at *ϵ* = 0, given Π(*ϵ*) defined by

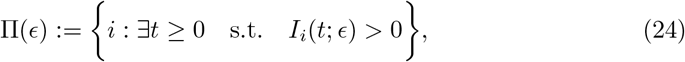

it will be assumed that Π(*ϵ*) = {1, …, *n*} for all *ϵ* > 0 (as any groups which never suffer any infections can be ignored) and that Π(0) = {2, …, *n*}. While this second condition may not be strictly necessary for the theorem to hold, it is unrestrictive, and ensures convergence - if this were not the case, then it would be possible that infection in some set of groups were seeded only by group 1. Thus, when *ϵ* = 0, these groups would suffer no infections, while for any *ϵ* > 0, they would have an epidemic of size independent (at leading order) of *ϵ*.

The final condition on the model is that the people in group 1 can be infected by other groups, and that vaccinated members of group 1 gain protection from this infection. That is, there is some *i* ∈ {1, …, *n*} such that

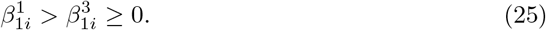

This is an important condition, as if people group 1 could only be infected by other members of group 1 the the total number of infections in group 1 would decay as *ϵ* → 0, meaning that it would no longer necessarily be optimal to vaccinate group 1 first (as most people in group 1 would not catch the disease anyway for small *ϵ*).

With these considerations, Theorem 1 can now be stated.

##### Theorem 1

*Suppose that for all ϵ* > 0,

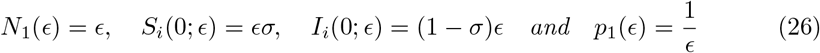

*for some σ* ∈ (0, 1). *Suppose that all other parameter values and initial conditions are independent of ϵ*.

*Consider any vaccination policy with uniformly bounded finite support given by* ***U*** (*t*; *ϵ*) *and suppose that there exists fixed α, τ, w* > 0 *such that*

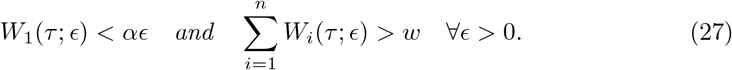

*Define a new policy*, ***Ũ***(*t*; *ϵ*), *given by*

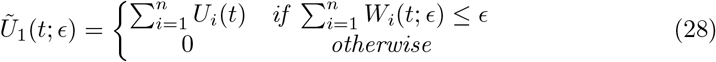

*and, for i* ≠ 1,

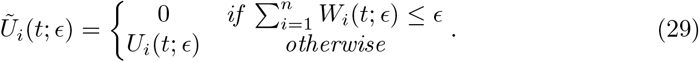

*Suppose that for each i* ∈ {1, …, *n*} *and t* ≥ 0,

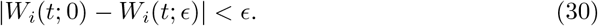

*Define*

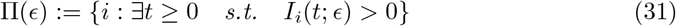

*and suppose that* Π(*ϵ*) = {1, …, *n*} *for any ϵ* > 0 *and that* Π(0) = {2, …, *n*}. *Finally, suppose that there exists a i* ∈ {2, …, *n*} *such that*

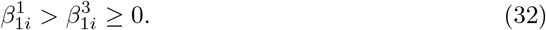

*Then, the policy* ***Ũ*** *is feasible and for sufficiently small ϵ*,

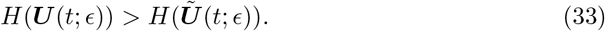

For the second theorem, it is helpful to note that, using the results in [24], if one defines

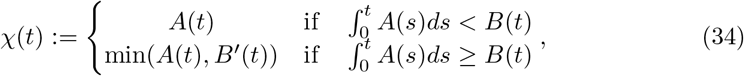

then (assuming that there is an optimal solution, and under mild smoothness conditions on ***U***, *A* and *B*) there must be an optimal solution satisfying

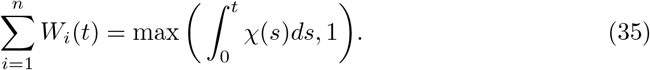

The following theorem then proves that the limiting optimal vaccination policy vaccinates the vulnerable group as quickly as possible. To reduce the length of the proof, it will be assumed that *σ* = 1, so that (in the small *ϵ* limit) all members of group 1 can be vaccinated before being infected.

##### Theorem 2

*With the definitions of Theorem 1, suppose additionally that*

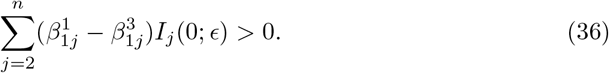

*That is, the initial difference between the infective force on vaccinated and unvaccinated members of the population is positive. Suppose further that*

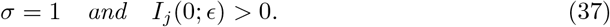

*Suppose an optimal vaccination policy for each ϵ is given by* ***Ū***(*t*; *ϵ*) *and suppose that* ***Ū***(*t*; *ϵ*) *has uniformly bounded finite support. Then, there exists an η depending only on α, τ, w and the model parameters such that, for any* ***U*** *satisfying the condition (27) as defined in Theorem 1*

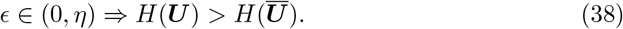

*Moreover, there is a sequence of optimal vaccination policies*, ***Ū***(*t*; *ϵ*), *which satisfies*

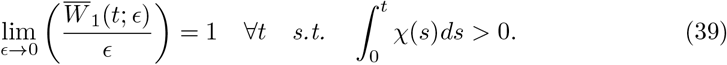

Theorems 1 and 2 are proved in 5.

#### 3.1.2 Examples

To illustrate these analytic results, consider a simple two-group example. Suppose that group 1 is small, vulnerable, and non-infectious, while group 2 is large, invulnerable and infectious. These groups could be interpreted as “old” and “young” respectively, although there is no specific physical situation being modelled here.

Suppose the transmission matrices are given by

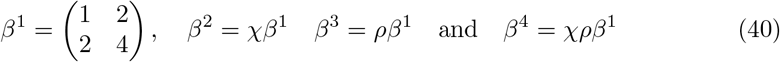

for some parameters *χ* and *ρ* which will be varied. This corresponds to the case of vaccination having (independently) an effectiveness *χ* at stopping people being infected and *ρ* at stopping infected people transmitting the disease. Moreover, suppose that

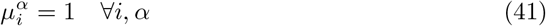

and

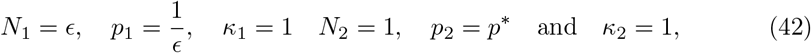

for some parameter *p*^*^ that will be varied. Finally, suppose that the initial conditions are

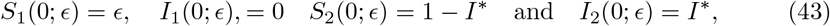

for some parameter *I*^*^ that will be varied, and that the vaccination constraints are given by

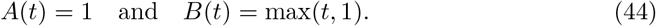

Consider therefore a vaccination policy where group 2, the infectious group, is vaccinated first (and hence, as *B*(∞) = *N*_2_, it is the only group that is vaccinated). That is,

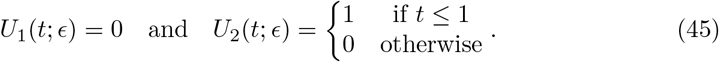

Hence, with ***Ũ*** defined as in Theorem 1, one has

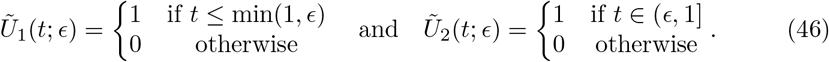

Fig. 1 shows a comparison of the objective values *H*(***U*** (*t*; *ϵ*)) and *H*(***Ũ***(*t*; *ϵ*)) for different values of *ϵ*. As expected, when *ϵ* = 1, vaccinating the more infectious group first is optimal (as they have the same vulnerability in this case), while for *ϵ* smaller than around 0.1, it becomes more effective to vaccinate the vulnerable group first, illustrating the results of Theorem 1.

**Fig 1.**
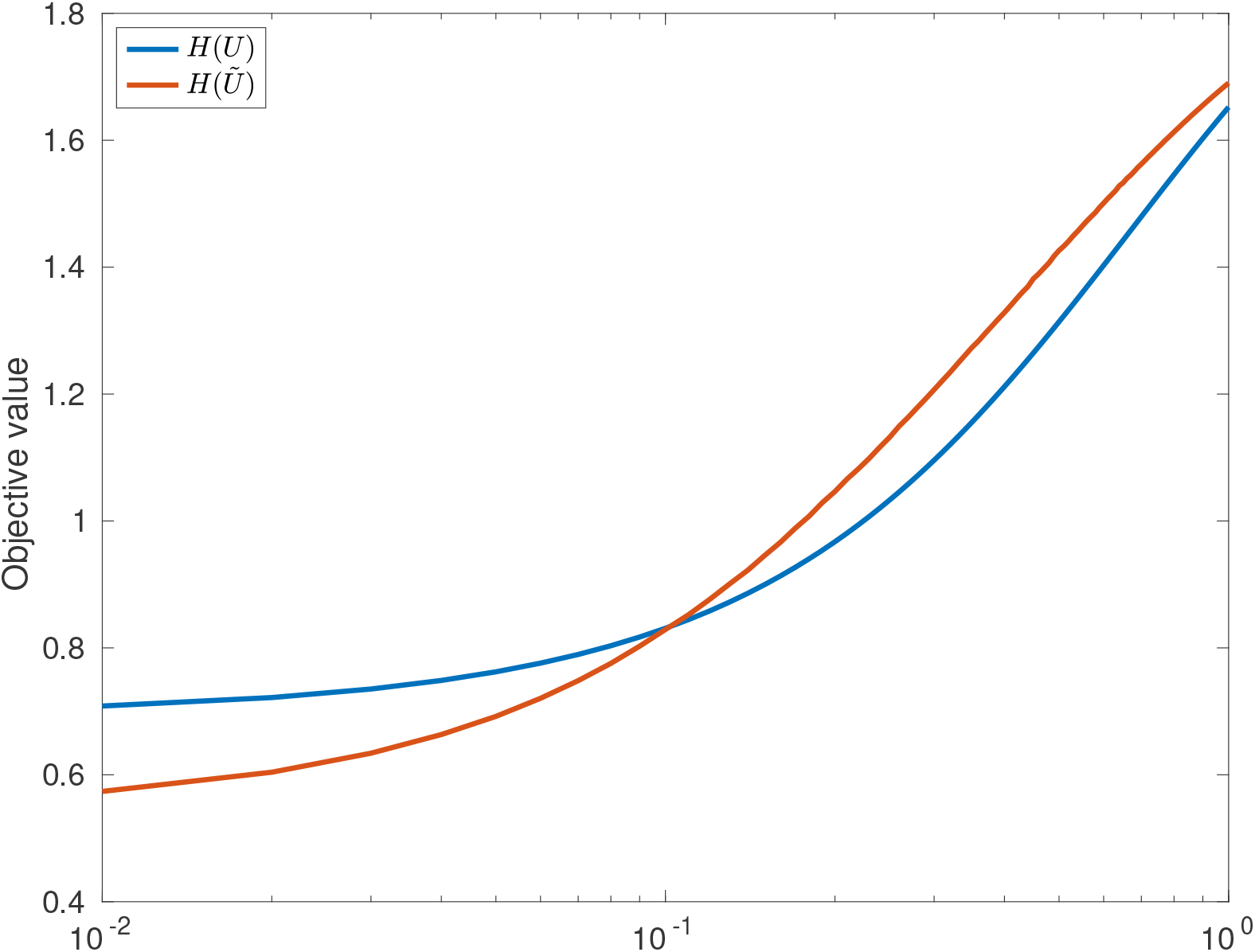
A comparison of the two vaccination policies, ***U*** (*t*; *ϵ*) (where the infectious group is vaccinated first) and ***Ũ***(*t*; *ϵ*) (where the vulnerable group is vaccinated first) for different values of *ϵ*. Note that here, *I*^*^ = 0.01, *χ* = *ρ* = 0.5 and *p*^*^ = 1.

It is useful to consider the approximate smallness of *ϵ* required in Theorem 1. That is, how small *ϵ* needs to be in order for ***Ũ***(*t*; *ϵ*) to be the better vaccination policy. To explore this, define, for each value of *I*^*^ and *p*^*^,

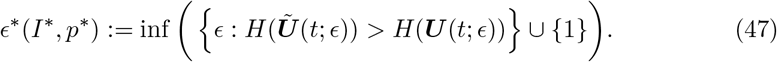

That is, *ϵ*^*^(*I*^*^, *p*^*^) is the smallest value of *ϵ* such that vaccinating group 2 first is better that the ***Ũ*** policy, with a cut-off value at 1 (as it is possible that for some parameter values, the ***Ũ*** policy is always better).

Fig. 2 shows the behaviour of *ϵ*^*^(*I*^*^, *p*^*^). As expected, *ϵ*^*^ is decreasing in *I*^*^ - this is because when there are fewer initial infectives, there is more time to vaccinate the infectious group before the epidemic has a chance to grow, reducing the peak of the epidemic. Moreover, *ϵ*^*^ is decreasing in *p*^*^, as higher values of *p*^*^ mean that the number of infections in group 2 is valued higher.

**Fig 2.**
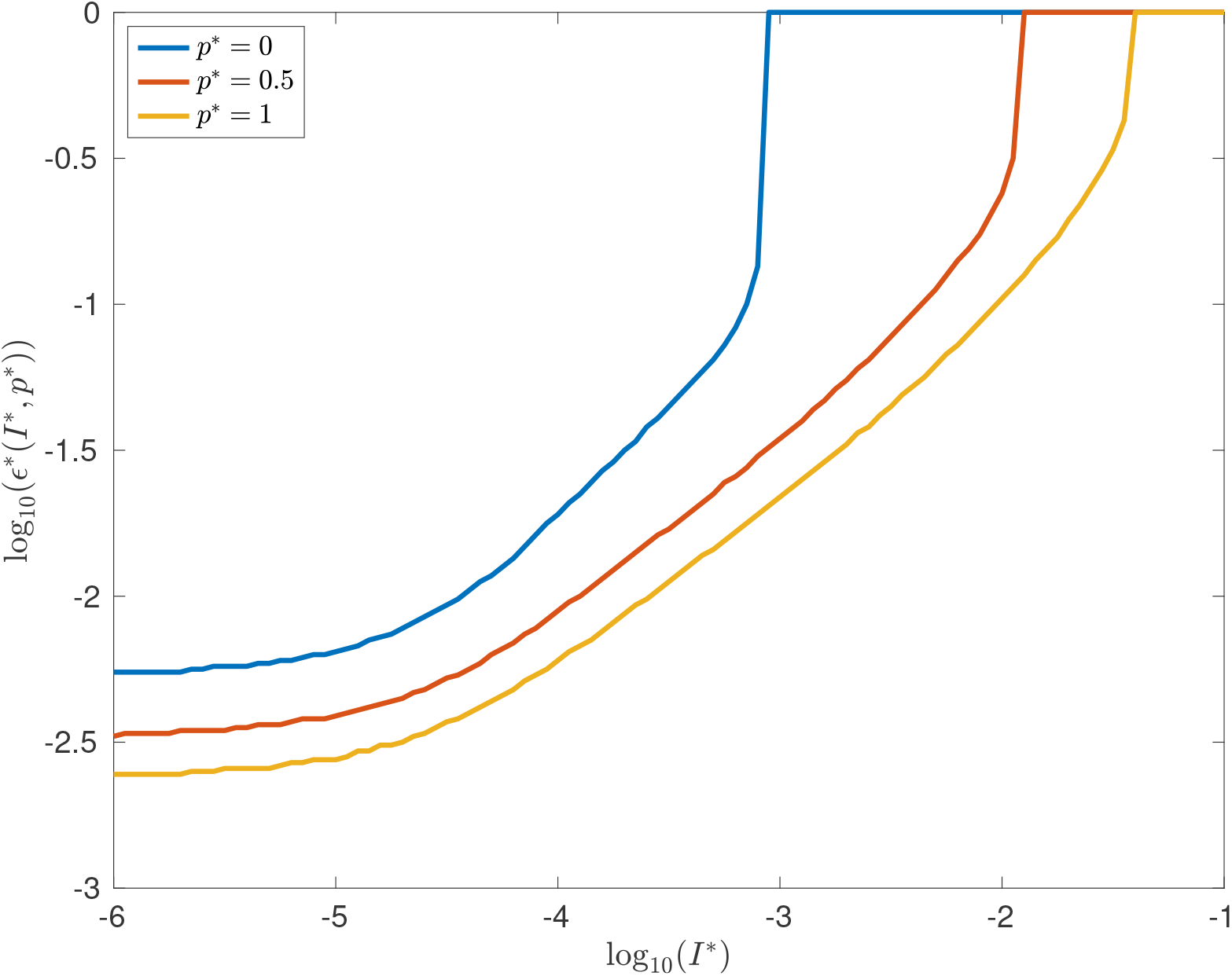
A plot of *ϵ*^*^(*I*^*^, *p*^*^), the highest value of *ϵ* for which ***U*** is a better vaccination policy that ***Ũ***. Note that *ϵ*^*^ is capped at 1, so that a value of 1 indicates that there were no values found of *ϵ*^*^ such that ***U*** was the better policy. Note that here, *χ* = *ρ* = 0.5.

Moreover, Fig. 2 suggests that, for each fixed *p*^*^, *ϵ*^*^ is uniformly bounded below for all *I*^*^. Indeed, this is expected as when *I*^*^ is very small, there are negligible infections within the interval *t* ∈ [0, 1] and so the vaccination policies ***U*** and ***Ũ*** are in effect being carried out in a completely uninfected population. As the *R*_0_ (that is, the initial growth rate of the disease) number of a fully vaccinated population (in this case) is greater than 1, *I*(*t*; *ϵ*) will reach an *O*(1) value regardless of the vaccination policy. Thus, while decreasing *I*^*^ will increase the time to reach this *O*(1) value, it will not significantly change the final infections in the epidemic, and hence *ϵ*^*^ should converge to a fixed value for small *I*^*^.

When the fully vaccinated population has an *R*_0_ lower than 1, the difference between ***U*** and ***Ũ*** is more distinct. Indeed, provided *I*^*^ is small enough for vaccination to be completed before many infections have occured, one would expect *O*(*I*^*^) infections in group 2 in either of the two vaccination policies (for sufficiently small *ϵ*), as in both policies, the size of the infected compartment will be decreasing after the vaccination has been completed. However, in the ***U*** case, one would expect *O*(*I*^*^*ϵ*) infections in total in group 1 (as there is an *O*(*I*^*^) infection force on a group of size *O*(*ϵ*) for *O*(1) time), while in the ***Ũ*** case, one would expect *O*(*I*^*^*ϵ*^2^) infections in total in group 1, as the population of this group is only of size *O*(*ϵ*) for *O*(*ϵ*) time. This behaviour is illustrated in Fig. 3, which shows that *ϵ*^*^ converges to significantly higher values than in Fig. 2 - indeed, in the case that *p*^*^ = 0, it appears that ***U*** is never optimal for any *ϵ* ≤ 1.

**Fig 3.**
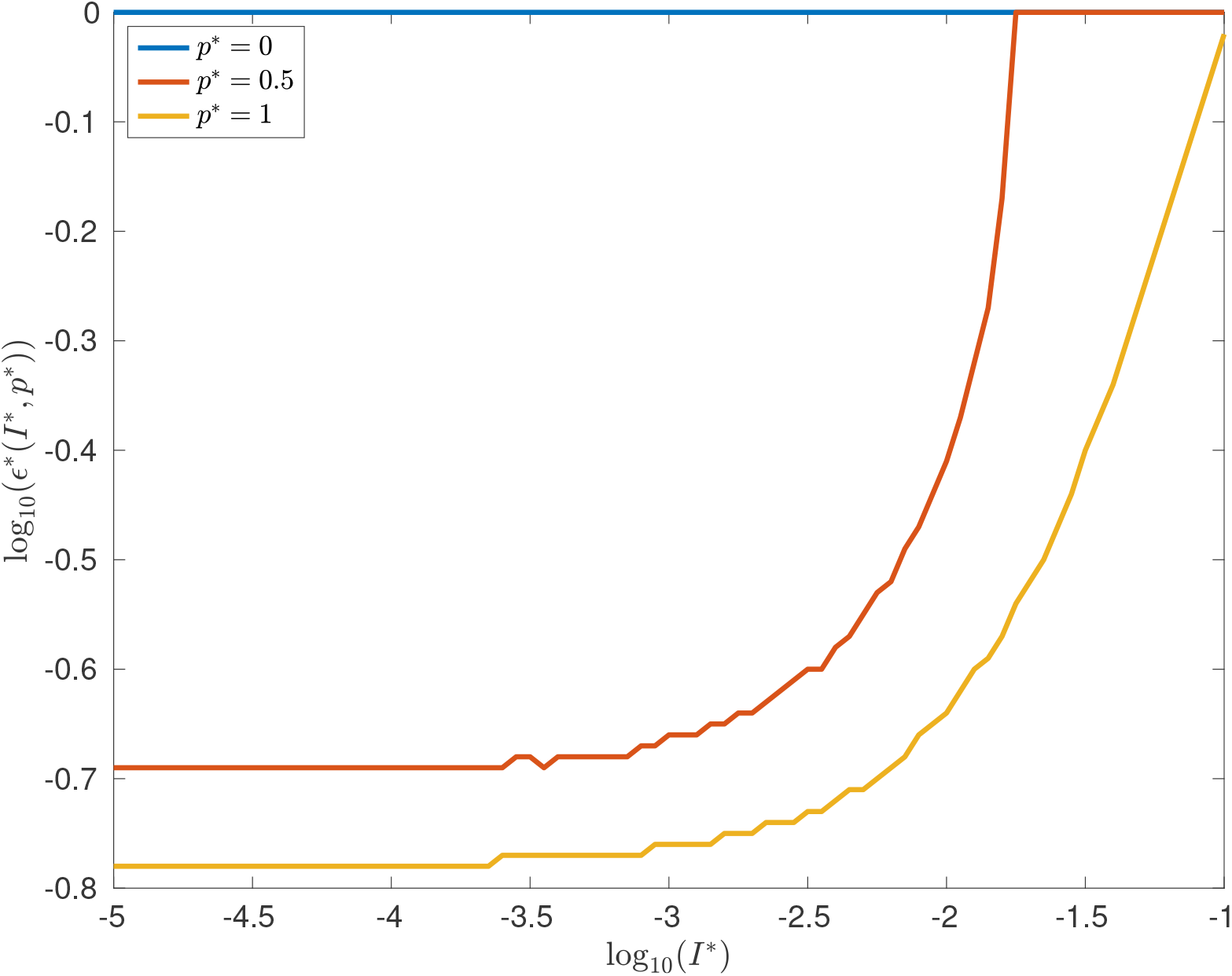
A plot of *ϵ*^*^(*I*^*^, *p*^*^), the highest value of *ϵ* for which ***U*** is a better vaccination policy that ***Ũ***, in the case of complete vaccination effectiveness (so *χ* = *ρ* = 0). Note that, because the values of the objective function are *O*(*I*^*^), there is some numerical instability which has caused some non-smoothness of the plot.

### 3.2. A small vaccination supply

In this section, the case of a small, immediately available vaccine supply will be considered. In this case, it will be possible to analytically derive the optimal vaccination policy (in the limit of small supply).

#### 3.2.1 Analytic results

To state the analytic result from this section, it is helpful to define

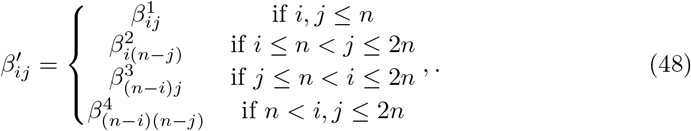

This large transmission matrix captures the dynamics of all 2*n* susceptible and infectious groups in the model (both vaccinated and unvaccinated). Indeed, after vaccination has been completed, there is no movement from *S*_*i*_ to 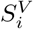 so *β*′ allows for the model to be considered as a 2n-group SIR model without vaccination. Thus, in particular, one can derive a simple final size relation for the total number of infections in the epidemic. Similarly, define

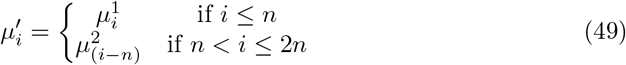

and

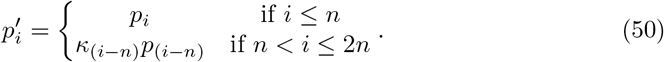

In this case of small supply, it is possible to effectively differentiate the final size of the epidemic with respect to the vaccination policy, and use the resultant linear approximation to form a simple knapsack problem for the optimal vaccination policy.

This will involve writing the objective in the form

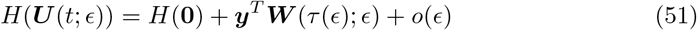

where ***W*** is the final vaccination amounts in each group. To define the gradient, ***y***, it is necessary to use the inverse of a matrix *M* given by

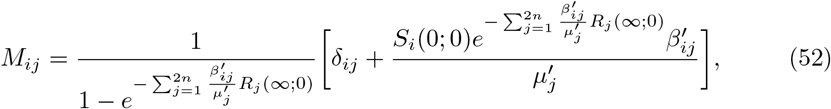

where as before, the variables *f*_*i*_(*t*; *η*) indicate the value of the relevant model variable at time *t*, given that the parameter *ϵ* is equal to *η*, and *δ*_*ij*_ is the Kronecker delta. Then, ***y*** is defined by

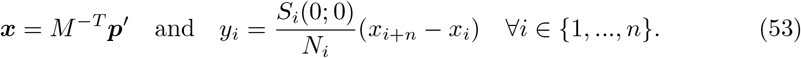

These definitions allow for the theorem to be stated.

##### Theorem 3

*Suppose that, for all ϵ* > 0

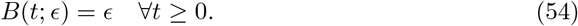

*and that all other parameter values and initial conditions are independent of ϵ. Suppose that A*(*t*) *is a continuous function with*

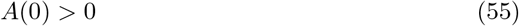

*and that the matrix M is invertible. For sufficiently small ϵ, define*

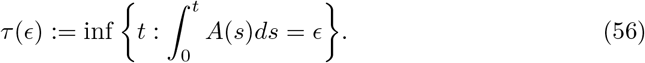

*Suppose that* ***U*** *satisfies the condition*

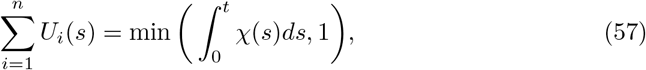

*where χ is defined in (35). Then, for sufficiently small ϵ, the objective function is given by*

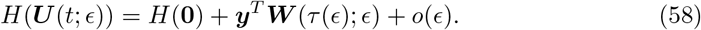

*Moreover, if there is a unique element of* ***y*** *equal to the minimum of* ***y*** *then the optimal vaccination policy (to leading order in ϵ) is uniquely given by*

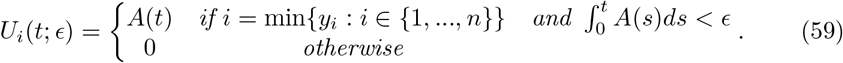

Theorem 3 is proved in 5.

#### 3.2.2 Vaccinating a homogeneous population

To illustrate the effectiveness of this approximation, consider first an example of a homogeneous population (so *n* = 1). Consider the case where *β*^1^ = *β, β*^2^ = *β*^3^ = 0.5*β* and *β*^4^ = 0.25*β* for some parameter *β* that will be varied. Suppose moreover that

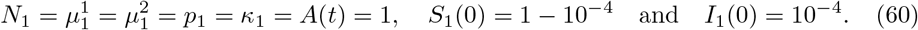

Finally, suppose *B*(*t*) = *ϵ* where *ϵ* will be varied.

Fig. 4 shows a comparison of the predicted and actual change in number of deaths, *ρ*_1_ for two values of *ϵ*. It illustrates that, even when *ϵ* = 0.1, a relatively large value, ***y*** gives a good approximation of the true value (found by simulation). Moreover, when *ϵ* = 0.01, the two lines are almost indistinguishable. This is useful validation for the approximation, as the correction term was simply proved to be *o*(*ϵ*) rather than, for example, *O*(*ϵ*^2^), and so it is encouraging that the predictions are so close.

**Fig 4.**
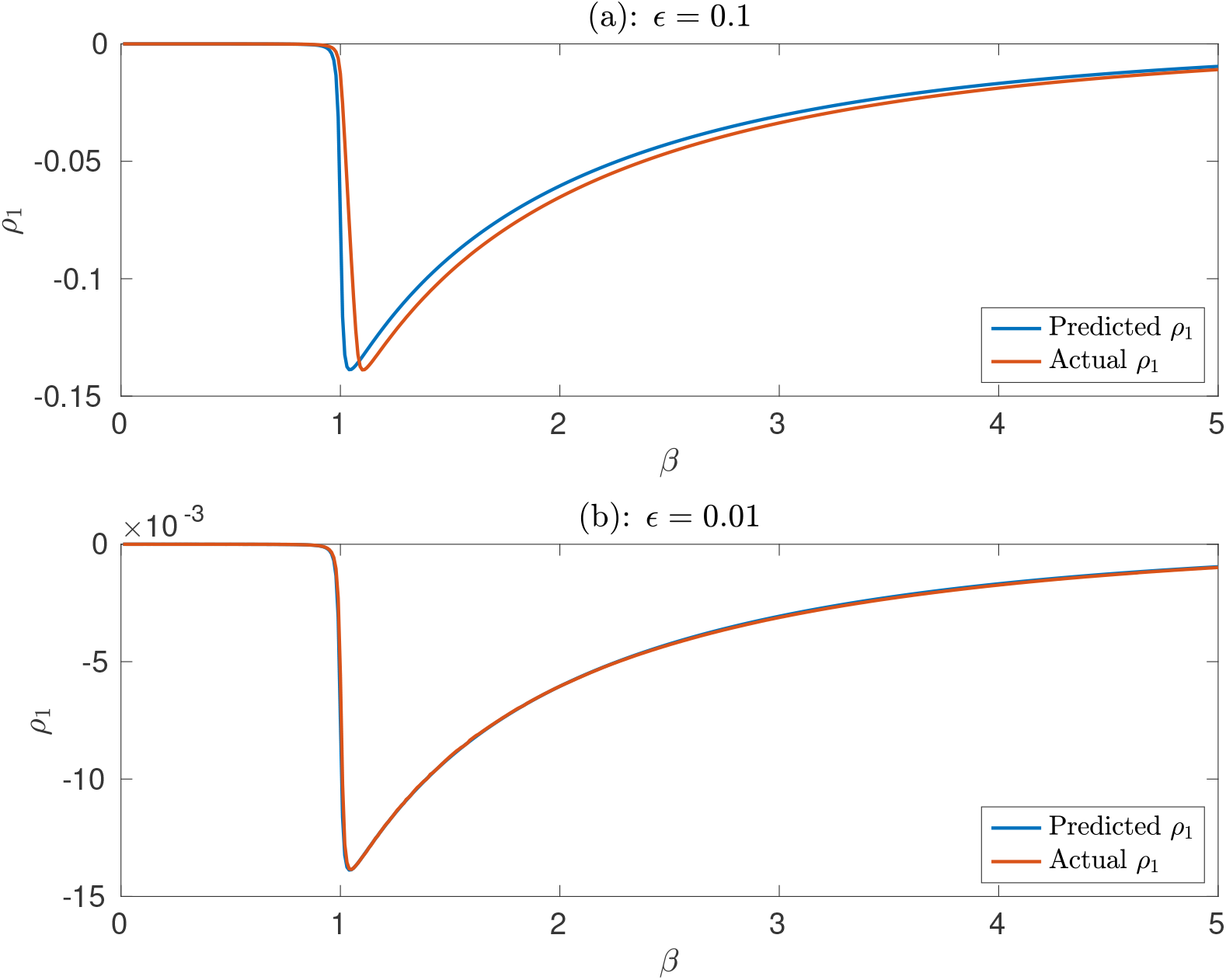
A comparison of the predicted and actual values of the change in deaths, *ρ*_1_, in the case of a homogeneous population for two different values of vaccination supply, *ϵ* and for different values of infectivity, *β*. Note the different scales on the two y axes.

An interesting property of Fig. 4 is that the value of *β* for which vaccination is most effective appears to be very close to *S*(0)*β* = 1 (as *S*(0) ~ 1). Note that here, as *μ* = 1, this is equal to the initial reproduction number of the disease. This has the perhaps surprising consequence that if one has a set of disconnected, equally vulnerable subgroups, a small vaccination supply should be assigned to a group with initial reproduction number close to 1, rather than giving it to the group with the highest value of *β* (that is, the most group with the most infectious individuals). This result is in line with the findings of [12] which showed that vaccinating less infectious groups can be more effective, and is an important consideration for vaccination policy planning.

#### 3.2.3 Application to age-structured populations

Consider assigning a small quantity of vaccinations to an age-structured population, using the example of the UK. The disease model has been estimated using the contact matrices **Λ** from [25], alongside population estimates ***N*** from [26]. As in [25], this gives a transmission matrix of

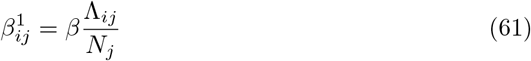

for some scalar parameter *β*. As in the previous section, it will be assumed that

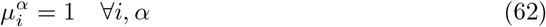

and

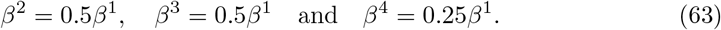

It will also be assumed that the initial infected population is small, so that, for each *i*

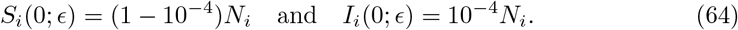

In the following examples, *β* will be chosen so that the disease-free next generation matrix of a completely unvaccinated population, given by

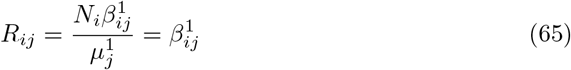

has a spectral radius (that is, largest eigenvalue) equal to 4. This sets the *R*_0_ number in the overall population to be 4. To illustrate the population structure, Fig. 5 shows a heatmap of the matrix *R*_*ij*_. This highlights the strongly assortative nature of the contacts (that is, members of a subgroup are most likely to be contacts with members of their own subgroup), while also showing that contacts are lower for older age groups.

**Fig 5.**
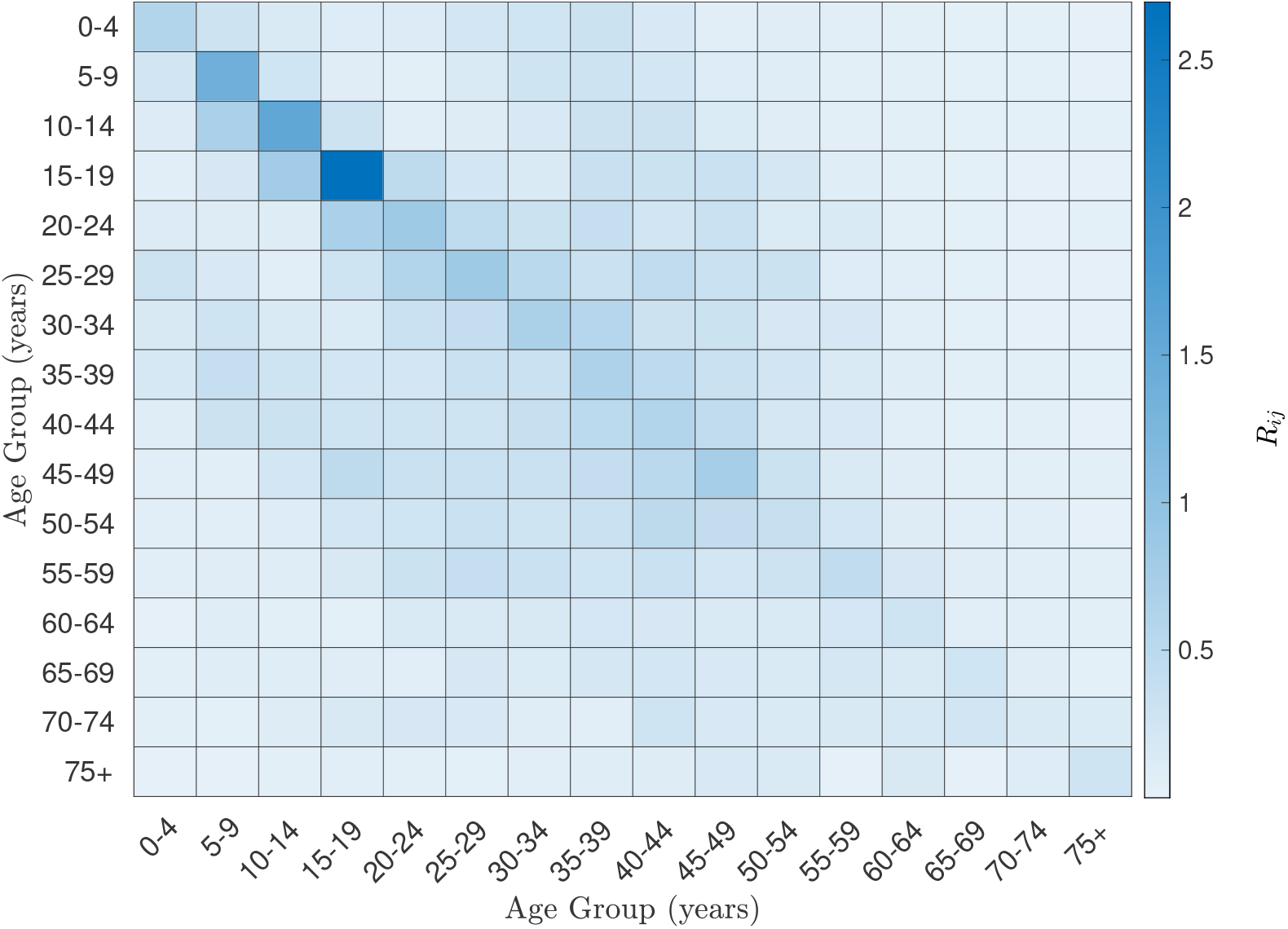
A heatmap of the next generation matrix for the age-structured UK population.

Now, two different age-dependent case-fatality ratios will be considered - uniform case-fatality and approximate COVID-19 case fatality, taken from [27]. In both cases, it will be assumed that vaccination reduces the case fatality ratio by 90% (following the results of [27] for the COVID-19 vaccines) so that *κ*_*i*_ = 0.1 for all *i*. However, it is worth emphasising that this model is simply based on real-world data to make this example more realistic, and does not seek to accurately model the COVID-19 pandemic.

Fig. 6 shows the effectiveness of vaccinating each age group in the two different cases, as a proportion of the optimal effectiveness. Note that here the proportion of effectiveness of assigning vaccine to group *i* is given by 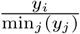, as each *y*_*j*_ is non-positive. It highlights that the significantly higher mortality rates for COVID-19 for the older age groups means that vaccinating them is much more effective than vaccinating the other age groups. This is an example of Theorems 1 and 2, as the oldest age group makes up a relatively small percentage (around 9%) of the population, but, if one scales *p* such that it has median value 1, the *p*_*i*_*N*_*i*_ value for the oldest age group is approximately 20, and so is definitely *O*(1) rather than *O*(*ϵ*).

**Fig 6.**
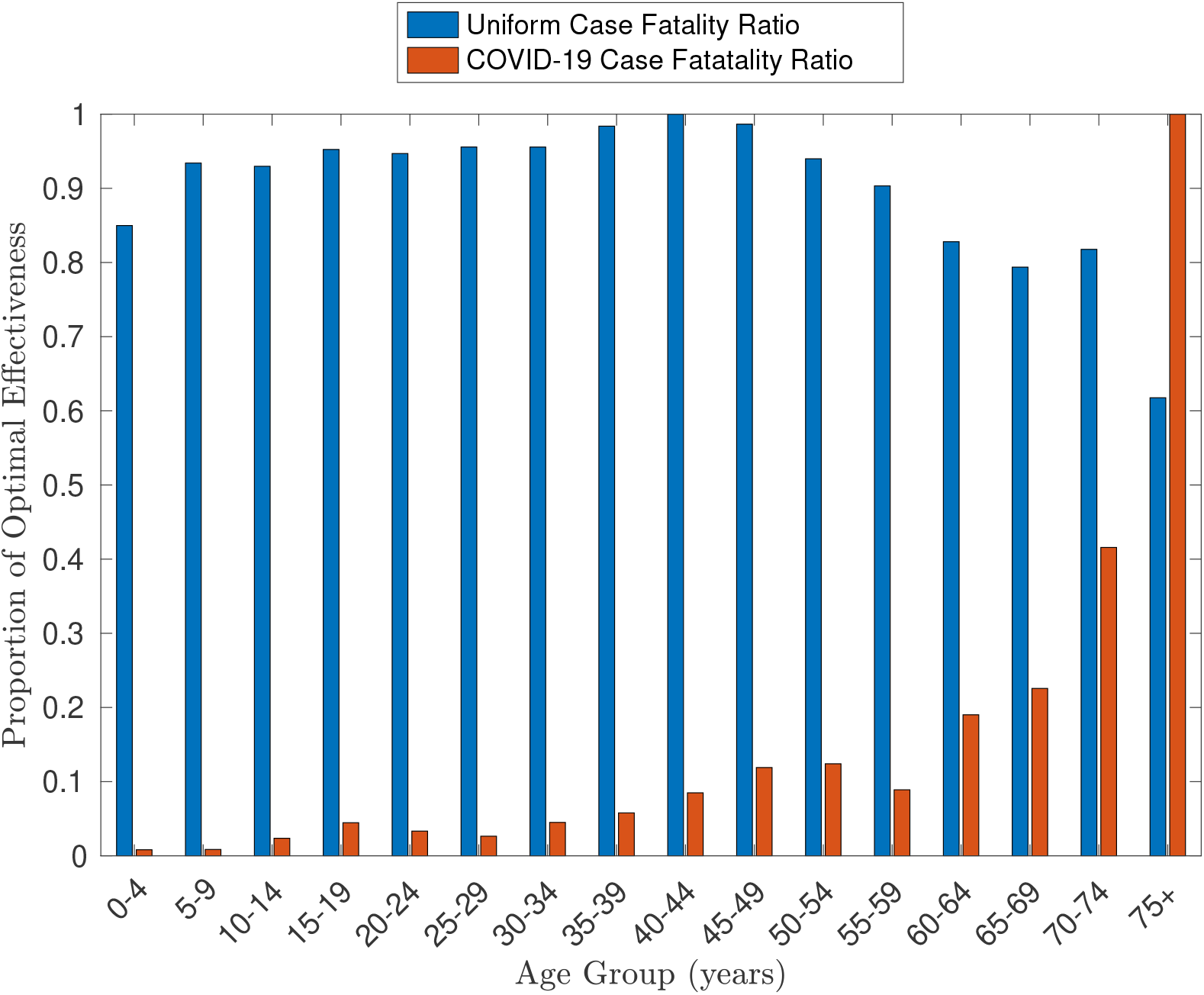
The effectiveness of assigning a the small quantity of vaccines to each age group as a proportion of the optimal effectiveness.

In the case of uniform mortality, it is perhaps surprising that the optimal age group to vaccinate is the 40-44 year olds. Indeed, from Fig. 5, it may seem that the 15-19 year old group would be the best group to vaccinate, as they have the highest overall transmission - that is, the maximum value of

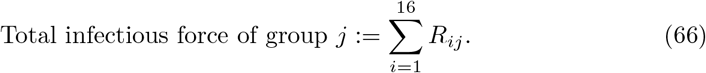

However, if instead, one considers

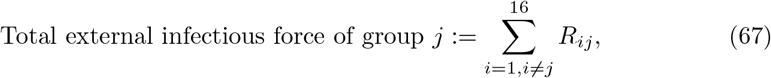

then it is the 35-39 and the 40-44 age groups which have the highest values. This can be considered in conjunction with the results of the previous subsection, which showed that vaccinating groups with *R*_0_ numbers close to 1 is optimal for disconnected populations. Indeed, the “secondary effect” of vaccinations (that is, the number of people who are not vaccinated, but are protected from the disease because of vaccines given to others) can be higher for groups with lower internal infectious force, particularly when their external infectious force is higher.

Finally, it is useful to again explore the range of values for *ϵ* for which **y** gives a good approximation of the true number of infections. As the minimum (scaled so that the total population size is 1) value of *N*_*i*_ is 0.0498 in this case, *ϵ* will be tested at 0.0498. The results of this are shown in Fig. 7, which again illustrates the effectiveness of this approximation. Indeed, the largest error across either case is of order 10^−4^, which in turn is of order *ϵ*^2^***y***. This suggests that the *o*(*ϵ*) correction term in Theorem 3 is significantly smaller than *ϵ*, which increases the usefulness of this approximation. However, further investigation is needed to determine whether this correction is of *O*(*ϵ*^2^***y***) for all parameter values.

**Fig 7.**
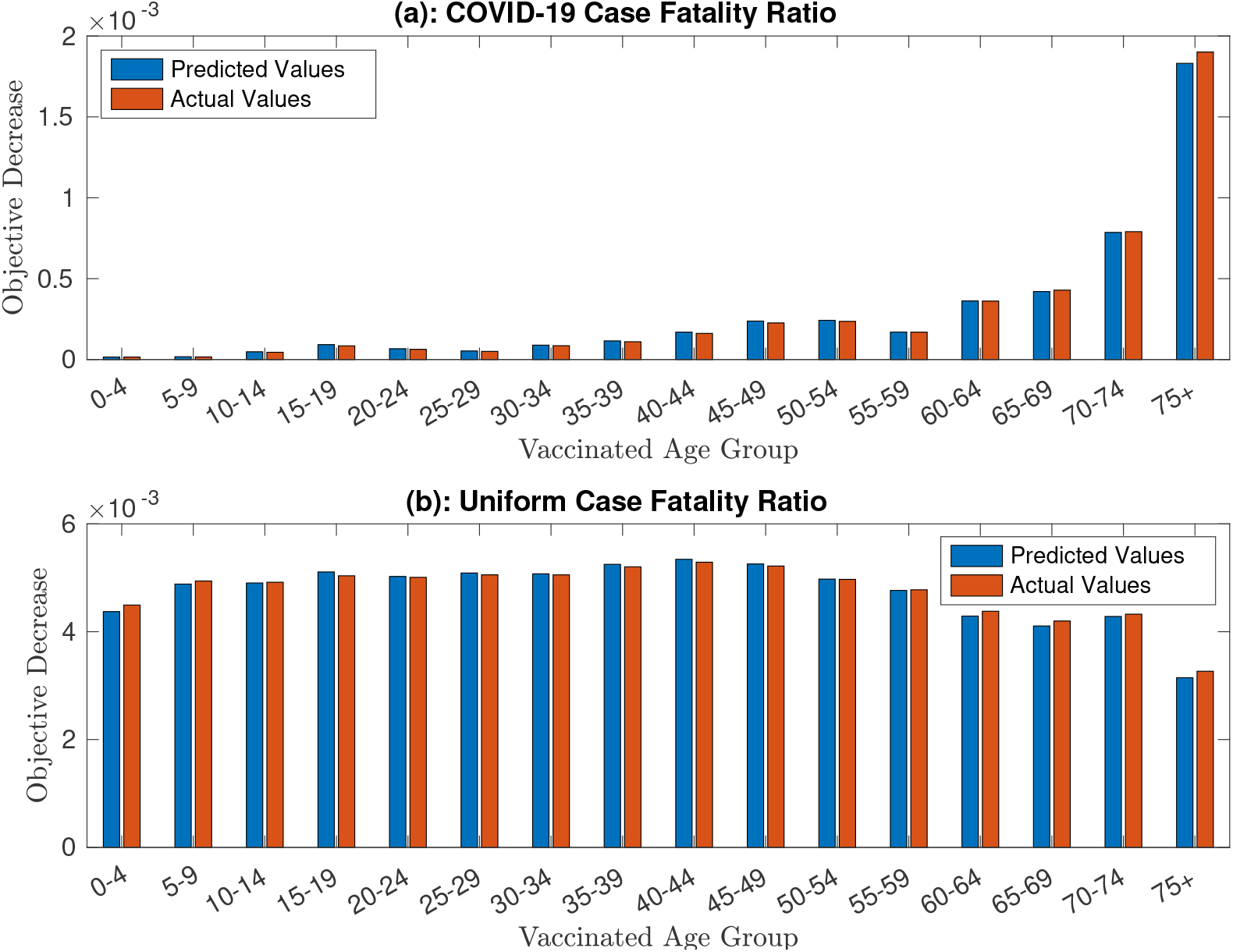
A comparison of the predicted and actual (from simulations) change in the objective function when vaccinating each individual group at *ϵ* = 0.0498. Both the cases of a COVID-19 case fatality ratio (in (a)) and a uniform case fatality ration (in (b)) are presented.

## 4 Discussion

This paper has shown two general principles for optimal vaccination policies by looking at the asymptotic behaviour of the optimal policy in the case of extreme parameters. Firstly, it has shown that small, vulnerable groups should in general be vaccinated first, regardless of the overall timetable of vaccination. This is an important result as it requires very little data on the population - merely the case fatality ratios and populations of the different subgroups - and in particular needs no forecasting of future transmission trends or vaccine supply.

The analytically derived results (in the limiting case) also show that the effect of vaccinating this small group far outweighs the effect of vaccinating any of the other groups. Indeed if the size of the vulnerable group is *O*(*ϵ*) and the case fatality ratio of the other groups is *O*(*ϵ*), then Theorem 1 shows that vaccinating the vulnerable group will lead to an *O*(*ϵ*) decrease in the number of fatalities, while vaccinating other groups will only decrease this by *O*(*ϵ*^2^). This provides strong evidence for the importance of sharing vaccines on a global scale to ensure that all of the most vulnerable can be vaccinated, as these are the vaccinations that will have by far the most significant effect on the global death toll from a pandemic.

It is important to note that this principle of vaccinating the most vulnerable group first may require *ϵ* to be very small, particularly when there are very few infectious people initially in the population, and when vaccination is not sufficient to eliminate the disease. Fig. 2 illustrated a scenario in which *ϵ* was required to be approximately 10^−3^ in order to guarantee that the vulnerable group was the best to vaccinate first. Thus, this result should be used with caution - it certainly does not imply that populations should always be vaccinated in order of vulnerability. Nevertheless, it seems likely that people that are extremely vulnerable, such as those with rare pre-existing conditions that make them especially susceptible to the disease, should always receive vaccines first.

The second principle derived in this paper was a linear approximation to the change in number of fatalities from a disease, which allows for the estimation of the optimal vaccination policy in the case of a small total supply. Again, this principle is flexible, applying for any underlying disease model, and provides a computationally cheap way of the approximating the optimal solution, even for large numbers of groups, as it merely requires the solution of a linear system involving the same number of parameters as the number of groups.

A useful feature of this approximation is that it appears to have high accuracy even for reasonably large values of the total supply, such as when 10% of the population can be vaccinated. Figs. 4 and 7 show that there is very little deviation between the predicted and actual values of the objective function and so suggest that this is a flexible and widely applicable method of approximation, even when the population contains a large number of subgroups. However, it would be helpful to strengthen the results of Theorem 3 to get a stronger bound on the error for small *ϵ* to ensure that this similarity holds for all models.

The results of the examples presented in Section 4 are also informative for vaccination policy. As shown in Fig. 4, in a completely homogeneous population, vaccination has the most effect when the reproduction number (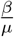 in this case) is slightly bigger than 1, with a steep decline in effectiveness for reproduction numbers below 1, and a more gradual decline for large reproduction numbers. This result allows one to consider the “vaccination leverage” of a population - that is, the effectiveness that a small quantity of vaccination can have - and shows that, even in the case of homogeneous case fatality ratios, vaccinating in order of infectiousness may be far from optimal, as it is much more difficult to reduce infections in a highly infectious population.

Indeed, a similar idea was shown to apply when the UK age structure was considered. In the case of uniform case fatality, the optimal group to assign a small amount of vaccinations to was the 40-44 age group which, as shown by Fig. 5, is not the most infectious group. This perhaps counter-intuitive results highlights the importance of mathematically justifying the principles one uses to decide on optimal vaccination policies, as “common-sense” arguments may in fact give false conclusions. Communicating such principles to governments and policy-makers will be crucial in future pandemics, particularly one with more homogeneous case fatality ratios where the optimal policy is not as intuitive as for diseases like COVID-19.

An important limitation of Theorem 3 is that the optimal policies for small vaccination supplies do not necessarily generalise to give the beginning of the optimal vaccination policy in the case of a much larger vaccination supply. Indeed, it is possible to have bifurcations in the optimal vaccination policy as the supply increases - for example, it can become possible to completely avoid an epidemic by vaccinating a large quantity of an infectious group. Thus, while the linear approximation can be a useful starting point when attempting to estimate the optimal strategy, it is important to consider alternatives when a large proportion of the population can be vaccinated.

This work could be extended by deriving more principles for extreme parameter values and investigating whether they generalise to realistic model parameters. By combining the existing results in this paper and others such as [12] with potential new ones, one could create an algorithm that creates good heuristics of optimal vaccination policies that could be used as starting points for accurately approximating the optimal policy for a general parameter set. This could have significant implications for the design of vaccination policies, as it would enable the optimisation problem to be estimated for very complex models, as the time taken to converge to an optimal solution would significantly decrease given good initial heuristics.

## 5 Summary and conclusions

The results of this paper are summarised below:

- For a sufficiently vulnerable, sufficiently small population in a multi-group SIR model, it is optimal to vaccinate this group first.
- For small overall vaccination supplies, the optimal vaccination problem can be well approximated by a simple knapsack problem.
- This linearisation appears to be a good approximation even for relatively large vaccination supplies (such as 10% of the population).
- This linearisation shows that, in the case of uniform case fatality, it is not necessarily optimal to vaccinate the most infectious group.

## Data Availability

The data and code underlying the results can be found at https://github.com/mpenn114/AsymptoticVaccination

## Nomenclature

inf: The infimum of a set
sup: The supremum of a set
*O*(*ϵ*): A function *f* (*t*; *ϵ*) which for sufficiently small *ϵ* is bounded by *Kϵ* for some constant *K*.
*o*(*ϵ*): A function *f* (*t*; *ϵ*) which satisfies lim_*ϵ*→0_(*f* (*t*; *ϵ*)*/ϵ*) = 0.

## Supporting information

**S1 Proofs. Proofs of Theorems 1,2 and 3**.

### S1 Proofs

This supplementary file contains the proofs of the theorems presented in the main text.

#### Proof of Theorem 1

##### Theorem 1

*Suppose that for all ϵ* > 0

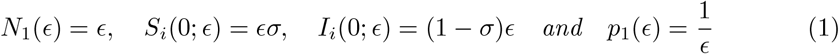

*Consider any vaccination policy given by* ***U*** (*t*; *ϵ*) *and suppose that there exists fixed α, τ, w* > 0 *such that*

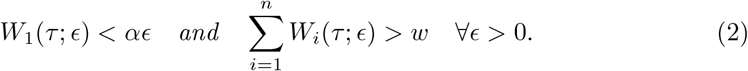

*Define a new policy* ***Ũ***(*t*; *ϵ*)

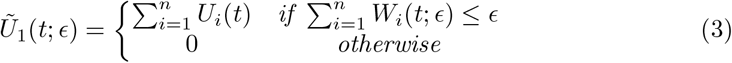

*and, for i* ≠ 1

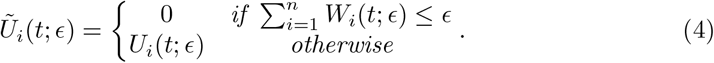

*Suppose that for each i* ∈ {1, …, *n*} *and t* ≥ 0,

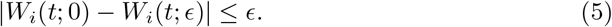

*Define*

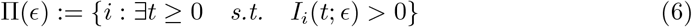

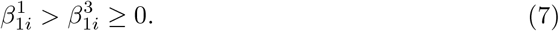

*Then, the policy* ***Ũ*** *is feasible and for sufficiently small ϵ*,

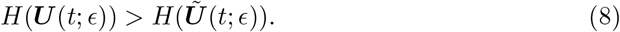

**Proof:** It is first important to prove that the ***Ũ*** is feasible. Firstly,

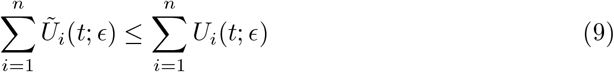

which, as ***U*** is feasible, means that the supply and rate constraints are satisfied. Moreover, as each *U*_*i*_(*t*; *ϵ*) ≥ 0,

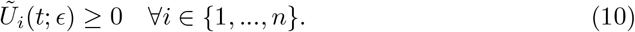

Also, for *i* ≠ 1,

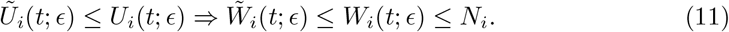

Finally, define

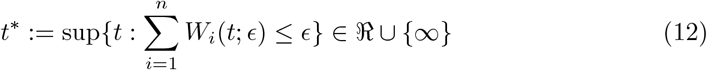

and then

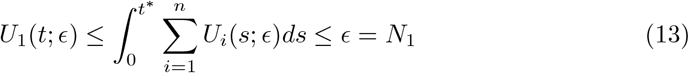

as required.

Define *S*_*i*_(*t*; *ϵ*) to be the number of susceptibles given the parameters *N*_1_(*ϵ*), *S*_1_(0; *ϵ*) and *I*_1_(0; *ϵ*) and the vaccination policy ***U*** (*t*; *ϵ*), and define 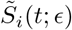 to be the number of susceptibles given the parameters *N*_1_(*ϵ*), *S*_1_(0; *ϵ*) and *I*_1_(0; *ϵ*) and the vaccination policy ***Ũ***(*t*; *ϵ*). Use similar definitions for the other variables in the model.

##### Proposition 1.1

###### Proposition 1.1

*For each t* ≥ 0 *and i* ∈ {1, …, *n*},

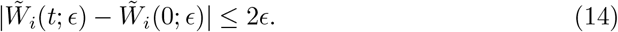

**Proof:** Firstly, note that

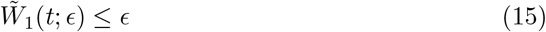

so

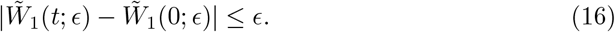

Now, suppose that *i* ≠ 1. Then, for each *ϵ, t* ≥ 0, with *t*^*^ defined as in (12),

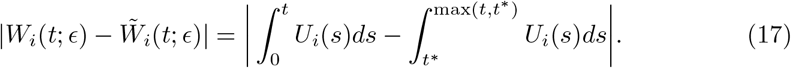

If *t* < *t*^*^, then

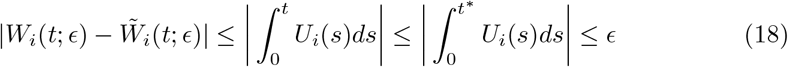

while if *t* ≥ *t*^*^, then,

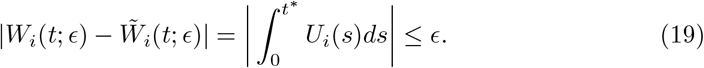

Thus, noting

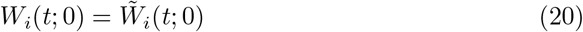

and using (5),

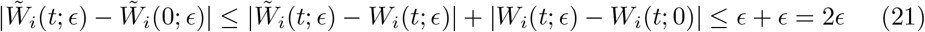

as required.

##### Proposition 1.2

###### Proposition 1.2

*Suppose that the U*_*i*_ *have uniformly bounded support for each ϵ* > 0. *Moreover, for each of the model variables, f*_*i*_, *suppose that*

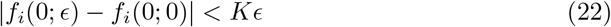

*for some constant K and that*

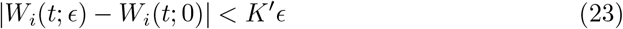

*for some constant K*^*′*^. *Finally, suppose all parameters are independent of ϵ with the exception that N*_1_(*ϵ*) = *ϵ. Then, for each δ* > 0, *there exists some* Δ > 0 *such that*

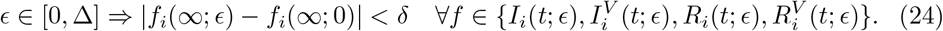

*Note that this holds both in the case of Theorem 1 (where N*_1_ → 0, Π(*ϵ*) = {1, …, *n*} *for ϵ* > 0 *and* Π(0) = 2, …, *n) or, in the case where each N*_*i*_ *is independent of ϵ (by adding a disconnected group of size ϵ)*.

**Proof:** Choose any *δ* > 0. Now, it is possible to write the system for ***I*** and ***I***^*V*^ in the form

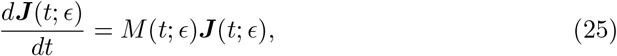

where *M* depends on the values of 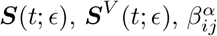 and 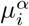 and

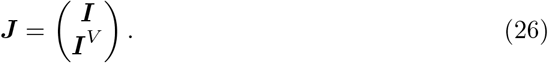

Hence, in particular, by using Proposition 1.1 and Lemma 4.7 for any fixed *t* ≥ 0,

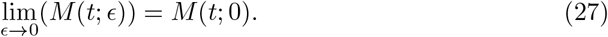

Moreover, if the support of each *U*_*i*_(*t*; *ϵ*) is bounded by *t*_*U*_ (which exists by assumption), then for *t* > *t*_*U*_, each *S*_*i*_(*t*; *ϵ*) and 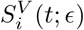 is non-increasing in *t* and so *M* (*t*; *ϵ*) is non-increasing. As it is bounded below, it therefore must converge to some matrix *M* (∞; *ϵ*), and, for *t* > *t*_*U*_,

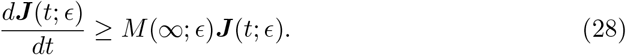

Hence, by Lemma 4.1,

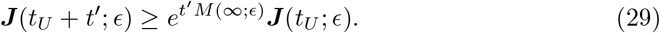

Moreover, by Lemma 4.3,

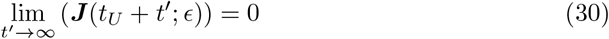

and hence (by non-negativity)

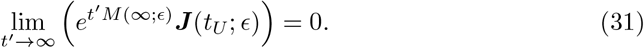

Now, define

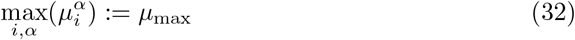

and then define

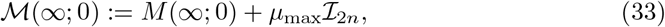

where ℐ_2*n*_ is the 2*n* by 2*n* identity matrix. Thus, in particular, ℳ(∞; 0) is non-negative and so

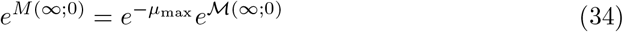

is non-negative as the exponential of a non-negative matrix is non-negative (as it is a weighted sum of powers of that matrix with positive weights). Thus, by Perron Frobenius theory, summarised in [1], there exists a real non-negative eigenvalue λ(∞; 0) (called the Perron eigenvalue) of *e*^*M*(∞;0)^ such that any other eigenvalues *ρ*(∞; 0) satisfy

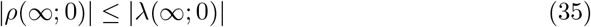

so, in particular

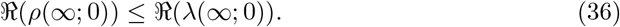

**Claim:** 0 < |λ(∞; 0)| < 1

**Proof:** Note that λ(∞; 0) > 0, as

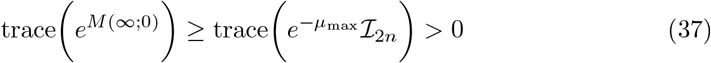

and thus, λ(∞; 0) ≠ 0.

From [1], there is a non-negative eigenvector, ***v***, with eigenvalue λ(∞; 0). Now, ***v*** must be an eigenvector of *M* (∞; 0) (as eigenvectors of a matrix and its exponential are the same). Thus, there is some λ^*^(∞; 0) such that

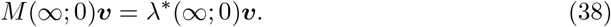

In particular, writing ***v*** = (*v*_1_, …, *v*_2*n*_)^*T*^

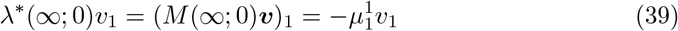

and thus, either 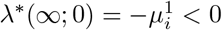 or *v*_1_ = 0. Suppose first that 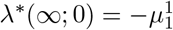.

Then, this means that (as the eigenvalues of *e*^*M*(∞;0)^ are the exponentials of the eigenvalues of *M* (∞; 0)),

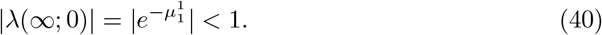

Similarly, *v*_*n*+1_ ≠ 0 implies that

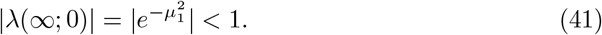

Thus, suppose for the remainder of the proof of this claim that *v*_1_ = *v*_*n*+1_ = 0. Now, for *i* ≤ *n*, the entries on the ith row of *M* (∞; 0) are given by

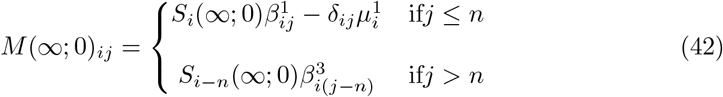

and for *i* > *n*, they are given by

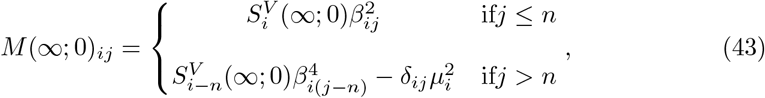

where *δ*_*ij*_ is the Kronecker delta.

Now, as Π(0) = {2, …, *n*}, by Lemma 4.5, it is necessary that

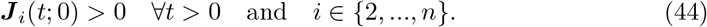

Moreover, if 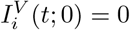 for some *t* > 0, then, by Lemma 4.8, as Π(0) = 2, …, *n*, it is necessary that

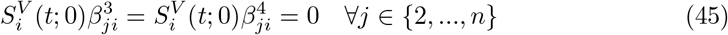

and so, if *t* ≥ *t*_*U*_, then this implies

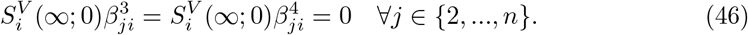

Thus, in this case, for *j* ∉ {1, *n* + 1}

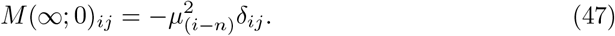

Therefore, suppose ***J*** _*i*_(*t*_*U*_; 0) = 0 for some *i* ∉ {1, *n* + 1} (and so, necessarily, *i* ∈ {*n* + 2, …, 2*n*}). Then,

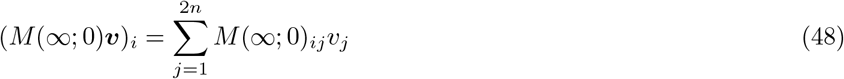

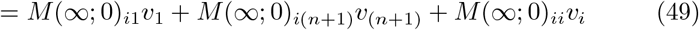

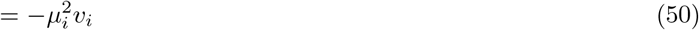

and so

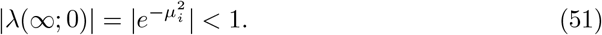

Consequently, this holds if any ***J***_*i*_(*t*_*U*_; 0) = 0. Conversely, suppose that ***J***_*i*_(*t*_*U*_; 0) ≠ 0 for all *i* ∉ 1, (*n* + 1). Then, there exists some *α* > 0 and some non-negative vector ***w*** such that

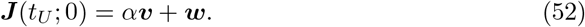

Therefore, for any positive integer *n*,

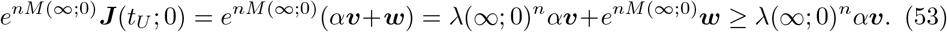

Now, ***v*** is an eigenvector so it has a non-zero component, which means that

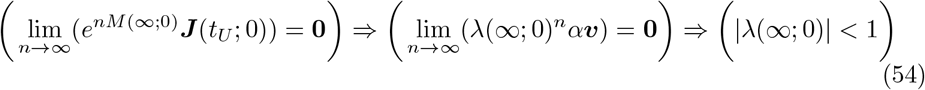

and so |λ(∞; 0)| < 1 holds in all cases, which finishes the proof of this claim.

**Claim:** *There exists some constant X independent of δ such that* 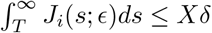

**Proof:** Now, the exponentials of the eigenvalues of *M* (∞; 0) are the eigenvalues of *e*^*M*(∞;0)^ which means that, if *η*(∞; 0) is an eigenvalue of *M* (∞; 0) then there exists some *κ* > 0 such that

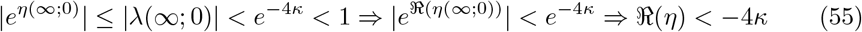

and so all eigenvalues of *M* (∞; 0) have strictly negative real part. Thus, by continuous dependence of eigenvalues on the matrix, as *M* (*t*; 0) converges to *M* (∞; 0) as *t* → ∞, there exists some *T* > *t*_*U*_ such that

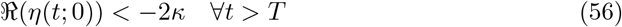

where *η*(*t*; 0) is an eigenvalue of *M* (*t*; 0). Now, fix *δ* > 0. From Lemma 4.3, by choosing *T* to be sufficiently large, one can assume that

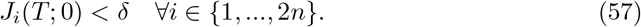

Moreover, there exists some Δ (which is dependent on *T*) such that

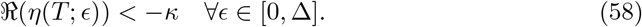

Now, similarly, by choosing Δ to be sufficiently small, one can assume that by Lemma 4.7

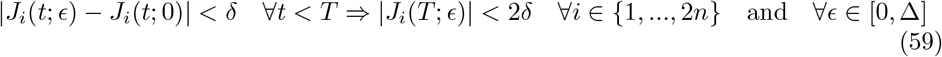

and

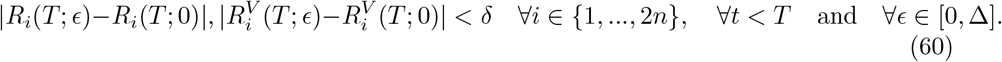

Now, for any *t* > 0,

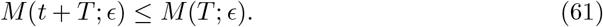

Thus, as the solution to the system

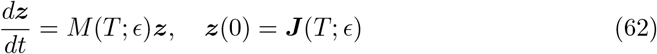

is

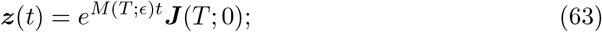

one has, by Lemma 4.1,

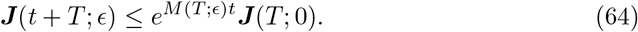

Now, noting that *M* (*T*; *ϵ*) is invertible as all its eigenvalues have strictly negative real part, for any *t* > 0

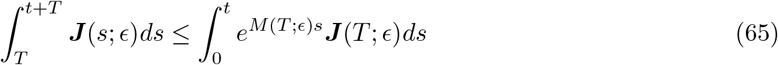

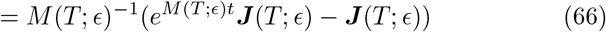

and so, taking *t* to ∞ and noting that all eigenvalues of *e*^*M*(*T*;*ϵ*)^ have real part less than 1 shows that

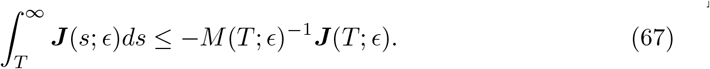

Now, each element of *M* (*t*; *ϵ*) is uniformly bounded (for any bounded range of *ϵ* and all *t* ≥ 0) as the parameters and variables are uniformly bounded. Thus, by expressing the inverse in terms of determinants of sub-matrices of *M* (*t*; *ϵ*) (each of which must be uniformly bounded as *M* (*t*; *ϵ*) is uniformly bounded) by Cramer’s rule [2], one can see that there exists a constant *M*^*^ such that for each *i* and *j*,

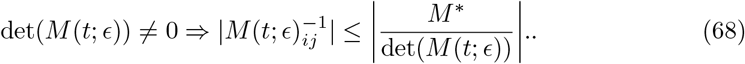

Note that

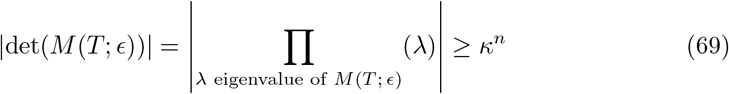

because all eigenvalues of *M* (*T*; *ϵ*) have real part at most −*κ* and hence modulus at least *κ*. Thus, there exists some constant *X* (independent of *δ*) such that for each *i* and *j*,

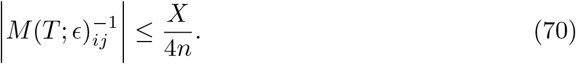

Thus, by the conditions on ***J*** (*T*; *ϵ*),

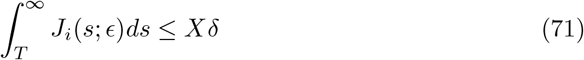

which completes the proof of this claim

As all the parameters and variables are uniformly bounded for all *ϵ*, there exists a constant *Y* (independent of *δ*) such that

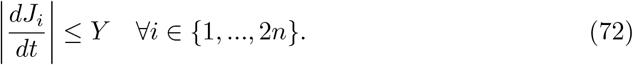

Now, suppose there exists some 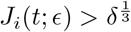 for *t* > *T* and *ϵ* ∈ [0, *η*_1_]. Then, by non-negativity of *J*_*i*_

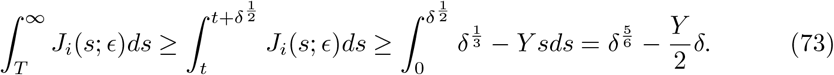

Thus, taking *δ* sufficiently small such that

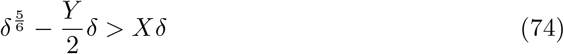

gives a contradiction. This means that, for each *i* ∈ {1, …, 2*n*}

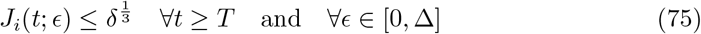

and hence, combining this with (59) (and assuming *δ* < 1 so 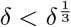) shows that

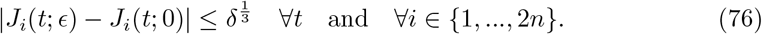

Moreover, by (71), for any *t* > 0

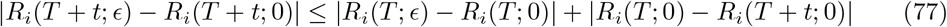

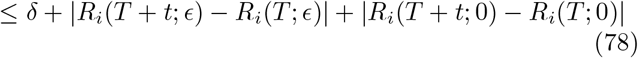

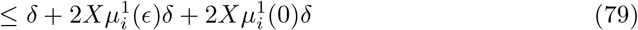

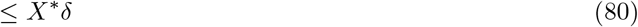

for some constant *X*^*^, alongside an identical result for 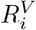. Combining this with (60) (and redefining *δ* → *δ*^3^), the result of the proposition is proved.

###### Theorem 1

Note that Proposition 1.2 also holds for the vaccination policies ***Ũ***(*t*; *ϵ*), using Proposition 1.1. Thus, one can define a function *δ*(*ϵ*) such that for all sufficiently small *ϵ*

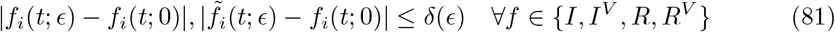

and

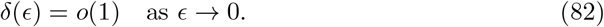

Then, using, for example

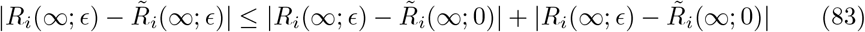

(ad 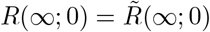) shows that

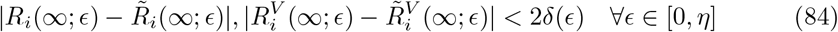

which means

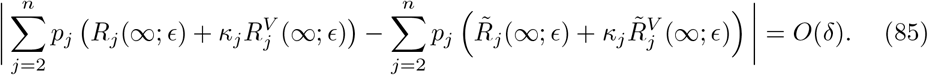

Thus, the aim of the remainder of the proof is to show that the leading order changes to *R*_1_(∞; *ϵ*) are of exactly *O*(*ϵ*), and so *p*_1_*R*_1_(∞; *ϵ*) changes by an *O*(1) amount, meaning these changes to the objective function will eventually dominate the other changes given in (85). This can be done by taking advantage of the fact that the quantities *f*_1_(*t*; *ϵ*) are small, and so a linearised version of the equations for group 1 can be used.

Before beginning this process, it is helpful to note the following. From (56) in the proof of Proposition 1.2, there exists some *T*^*^ > *t*_*U*_ independent of *δ* and *ϵ* such that

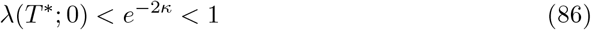

where λ(*T*^*^; 0) is the (necessarily real and non-negative) Perron eigenvalue of *e*^*M*(*T*\*;0)^ (and is the exponential of the *η*(∞; 0) referenced in (56)). Moreover, by the continuity of eigenvalues on the entries of the matrix, there exists some Δ > 0 such that the analogously defined λ(*T*^*^; *ϵ*) also satisfies

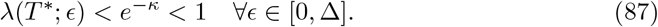

Now, note that, for *t* ≥ *T*^*^ > *t*_*U*_, the matrix *M* (*t*; *ϵ*) and hence the matrix *e*^*M*(*t*;*ϵ*)^ is non-increasing. Thus, as *e*^*M*(*t*;*ϵ*)^ is non-negative (as proved in Proposition 1.2), it is necessary from Perron Frobenius theory [1] that its Perron eigenvalue, λ(*t*; *ϵ*) satisfies

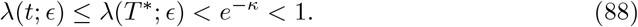

Then, following the method used to derive (67), one has, for any *t* ≥ *T*^*^

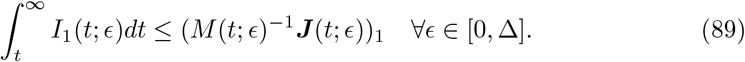

This is exactly the same equation as (67), except that here, *T*^*^ is independent of *δ* (as no conditions on ***J*** (*T*; 0) are assumed). Now, note that

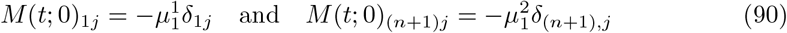

where here *δ*_*ij*_ is the Kronecker delta. This means that, for any vector ***y***, the equation

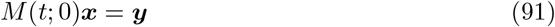

must satisfy

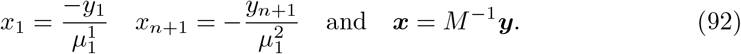

Thus, in particular

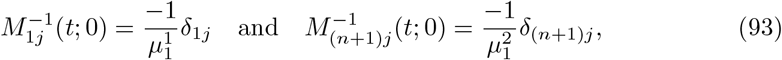

where here *δ*_*ij*_ denotes the Kronecker delta. Now, note that, as the inverse of a matrix is a rational function of its entries,

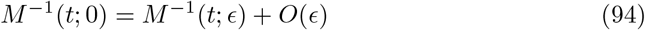

and hence

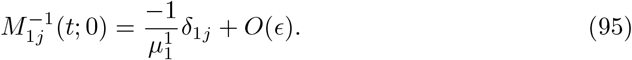

Moreover, defining

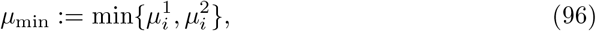

there must exist a 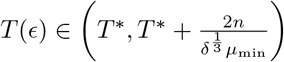 such that for each *i*,

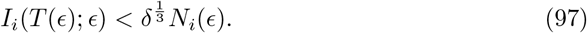

Otherwise,

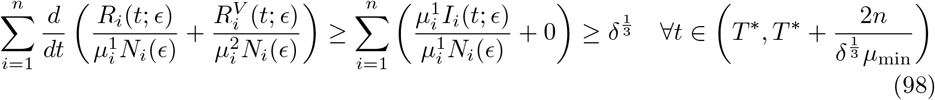

and integrating this between *T*^*^ and 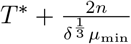 gives

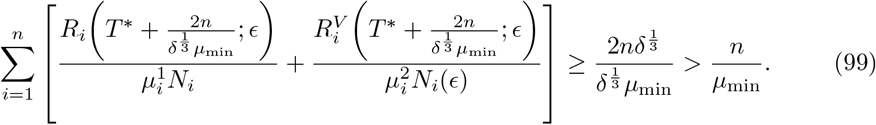

Thus, as 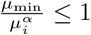 for each *i* and *α*,

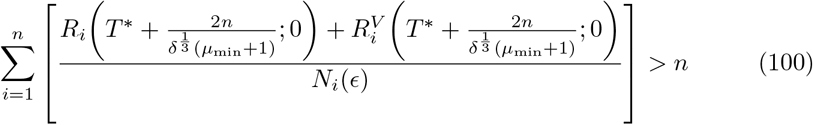

which means, for some *i*

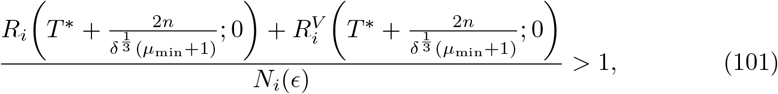

which is a contradiction as the total population size in group *i* cannot exceed *N*_*i*_(*ϵ*) by definition of *N*_*i*_(*ϵ*). Thus, for each *ϵ* ∈ [0, Δ],

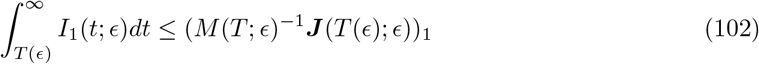

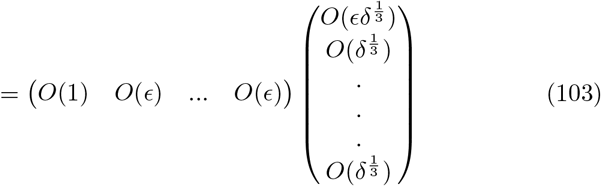

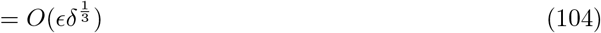

while similarly

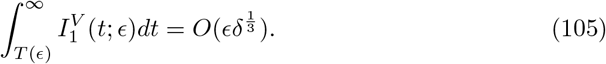

Moreover

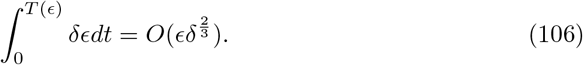

These results allow for the linearisation to be carried out. To reduce notation, define

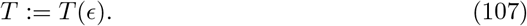

Now, to begin the linearisation, define

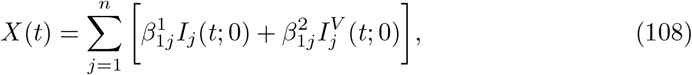

which is the leading order infective force on group 1. By Proposition 1.2,

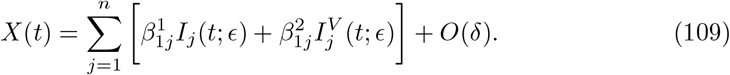

Then, as *S*_1_(*t*; *ϵ*) ≤ *ϵ*,

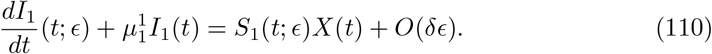

Now, note that

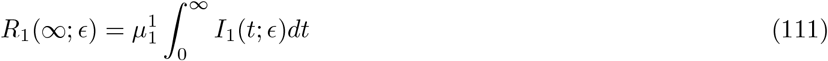

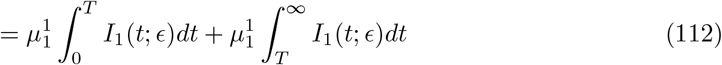

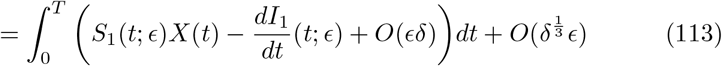

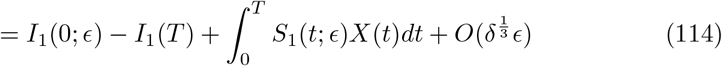

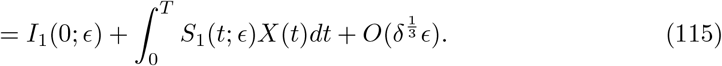

Now, the equations for *I*^*V*^ are of the same form, but with *S*^*V*^ in place of *S* and a different leading order infection function *Y* (*t*) given by

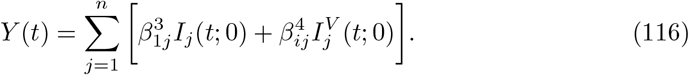

Thus, an analogous derivation (noting that *I*^*V*^ (0; *ϵ*) = 0) shows that

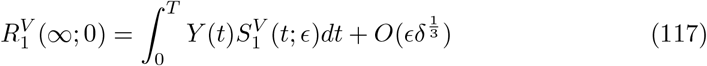

while analogous results hold for 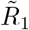 and 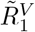 (with 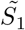 and 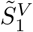 in place of *S*_1_ and 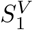). Now, note that

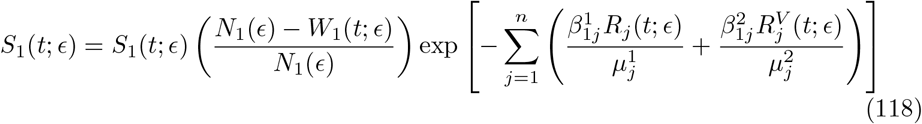

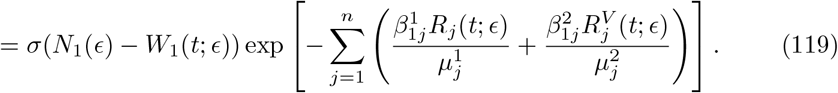

Define

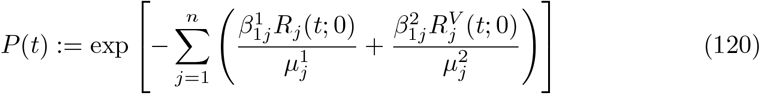

and then, note that by Proposition 1.2

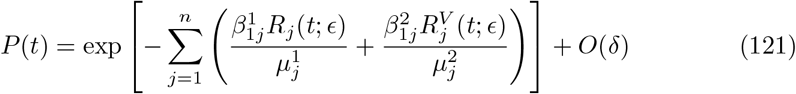

which means (as (*N*_1_(*ϵ*) − *W*_1_(*t*; *ϵ*)) ≤ *ϵ* and *σ* < 1)

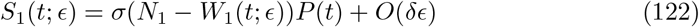

with an identical result for 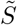. It is helpful to note for later that, as 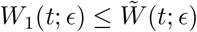, this means that

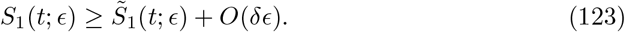

Now, this means

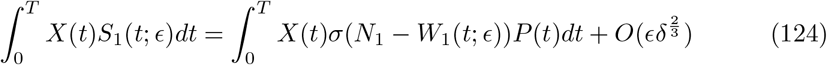

and so

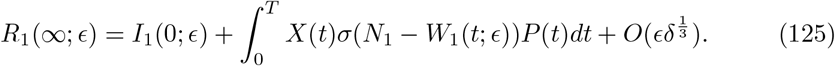

Now, note that

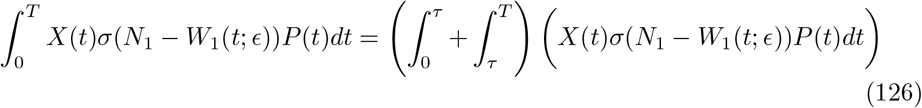

and that, as 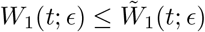,

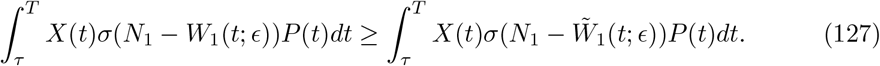

Now, define *z*(*ϵ*) to be

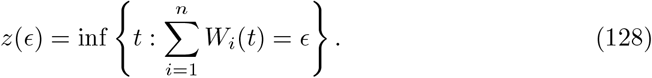

Note that, for *ϵ* < *w, z* exists and is bounded above by *τ* as

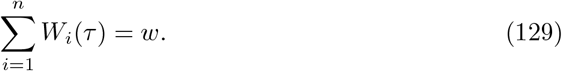

Now, define a fixed value

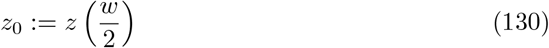

so that, by continuity of *W, z*_0_ < *τ* (and is independent of *ϵ*). Suppose that 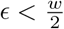 (which will be assumed for the rest of the proof). Note that

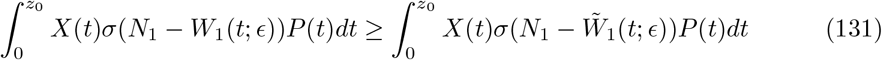

and that

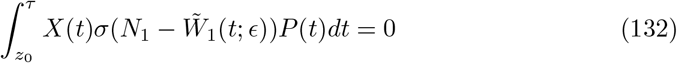

as 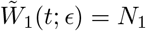 for all *t* > *z*(*ϵ*). Moreover, by (2)

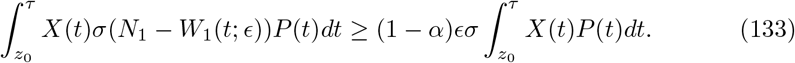

Now, note that *P* (*t*) is strictly positive for *t* > 0 as it is an exponential, while, as *β*_1*j*_ > 0 for some *j* ≠ 1,

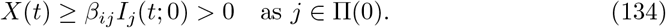

Thus,

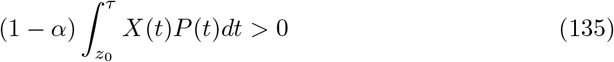

and this is independent of *ϵ*. This means that

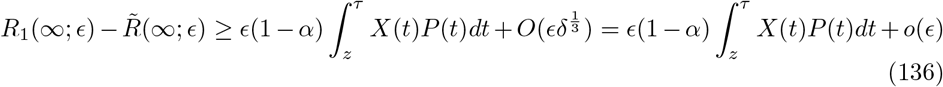

and so the leading order change in *R*_1_(∞; *ϵ*) is indeed of order exactly *ϵ*.

Now, it is important to check the leading order change in 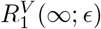. Note that, as *S*_1_(*t*; *ϵ*) and 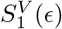 are at most *ϵ*,

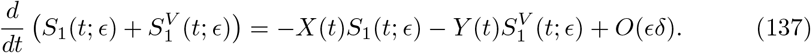

Using (122), this can be written as

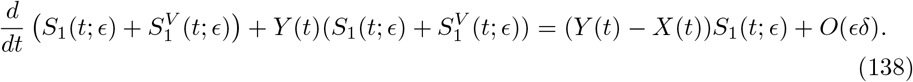

This equation can be integrated by defining

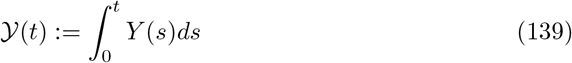

so that

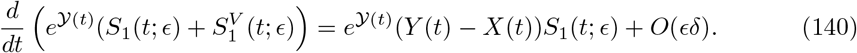

Thus, for any *t* ≤ *T*

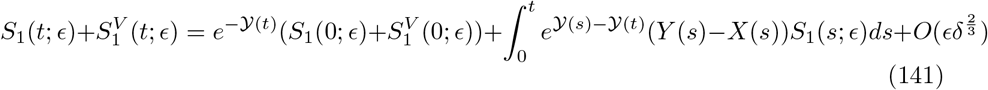

which means that

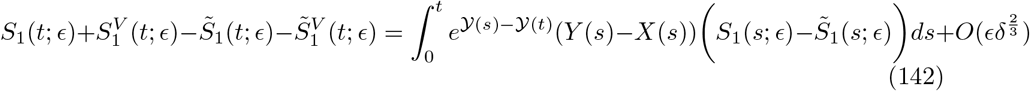

Thus,

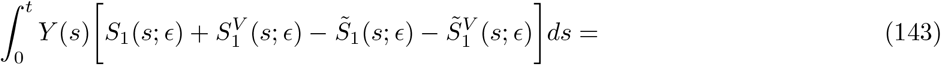

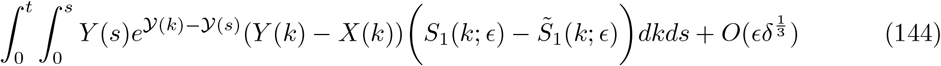

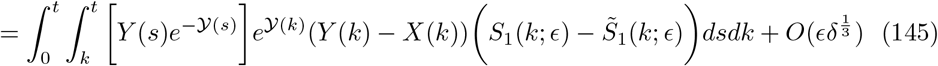

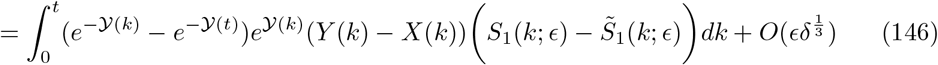

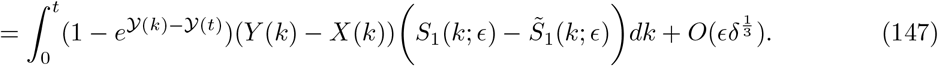

Now, note that, as *𝒴* is non-decreasing, and non-negative

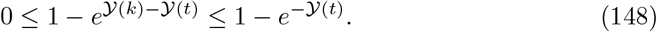

Moreover, one has

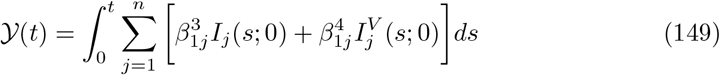

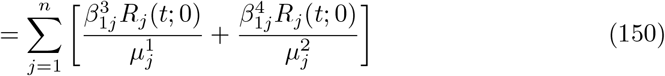

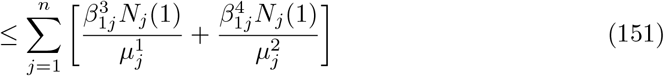

and so *𝒴*(*t*) is bounded above by some constant (for *ϵ* ≤ 1). This in turn means that there exists some *𝒴*\* such that

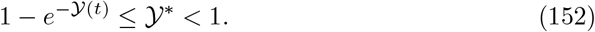

Thus, as *Y* (*t*) − *X*(*t*) ≤ 0 and 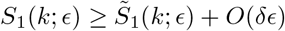, for any *k* ≤ *t*

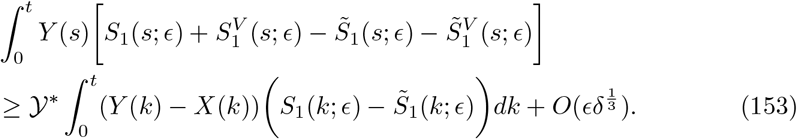

Now, adding the inequalities (115) and (117) together gives

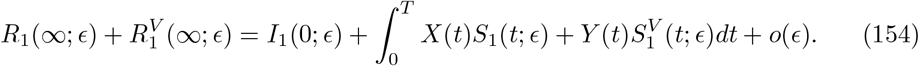

Note that

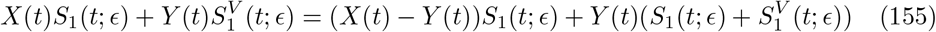

and hence

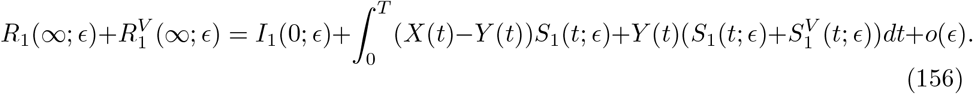

This means that

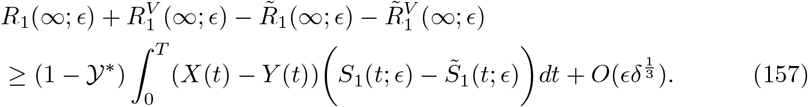

Now, as there is some *i* ≠ 1 such that

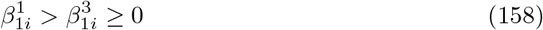

and (as *i* ≠ 1), *i* ∈ Π(0) which means that

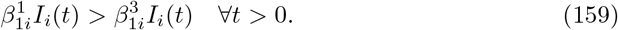

This means that *X*(*t*) > *Y* (*t*) for all *t* > 0 and hence

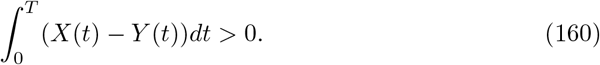

Thus, following the arguments from before, one can see that

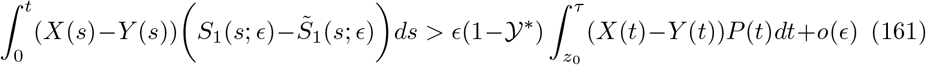

where the leading order term is positive as required (as *P* (*t*) is positive). Hence, from (157)

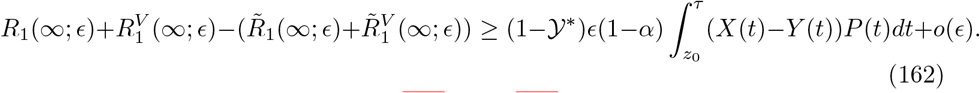

Thus, for any *κ*_1_ ∈ [0, 1], combining (136) and (162)

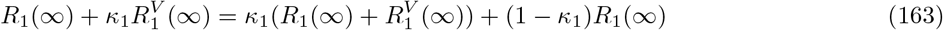

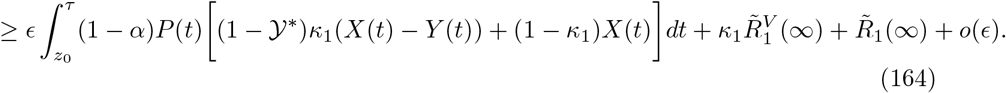

Thus, recalling (85) and that 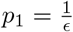

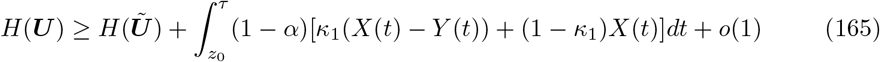

for some constant *K*. Moreover, for sufficiently small *ϵ*,

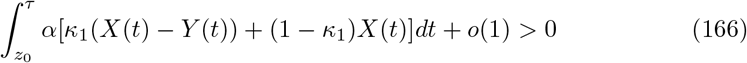

and hence

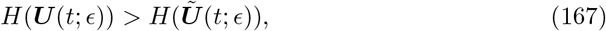

as required.

### 2 Proof of Theorem 2

Recall from the main text that, using the results in [3], if one defines

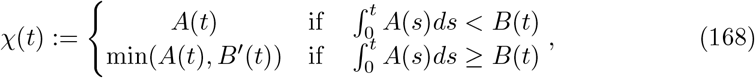

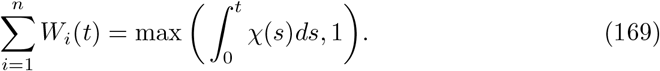

#### Theorem 2

*With the definitions of Theorem 1, suppose additionally that*

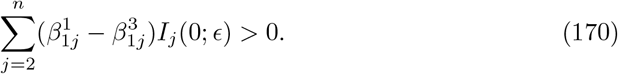

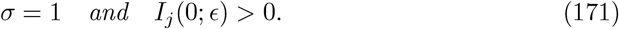

*Suppose an optimal vaccination policy for each ϵ is given by* ***Ū***(*t*; *ϵ*) *and suppose that* ***Ū***(*t*; *ϵ*) *has uniformly bounded finite support. Then, there exists an η depending only on α, τ, w and the model parameters such that, for any* ***U*** *satisfying the condition (2) as defined in Theorem 1*

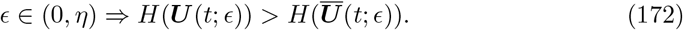

*Moreover, there is a sequence of optimal vaccination policies* ***Ū***(*t*; *ϵ*) *satisfying*

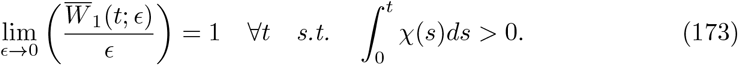

To make things clearer in the course of this proof, note that *H* will be written as

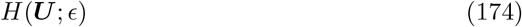

where the *ϵ* refers to the size of the population *N*_1_ under consideration.

#### Proposition 2.1

It remains to show that, for sufficiently small *ϵ* and fixed *α, τ* and *w*, there is no ***U*** satisfying the conditions (2) that is the optimal vaccination policy. To do this, the following proposition is required.

##### Proposition 2.1

*Suppose that I*_1_(0; *ϵ*) = 0 *for all ϵ. Consider, for ϵ* ≤ 1 *any bounded vaccination policy* ***U*** (*t*; *ϵ*) *given by*

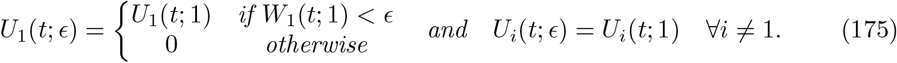

*Then, if H*(***U*** (*t*; *ϵ*); *ϵ*) *is the value of the objective function for a given value of ϵ*,

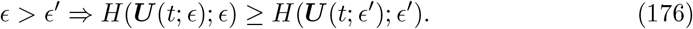

**Proof:** Fix *ϵ* and *ϵ*^*′*^ such that *ϵ* > *ϵ*^*′*^. Define the vaccination policy ***U*** ^*^(*t*, Δ; *ϵ*) to be

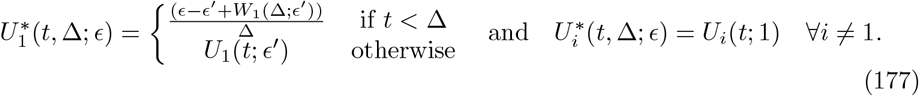

Then, in particular

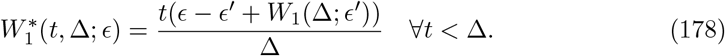

Now, as ***U*** (*t*; *ϵ*) is bounded by some *M*, it is necessary that *W*_1_(*t*; *ϵ*) is bounded above by *tM*. Conversely, 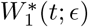 is bounded below by 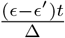 for *t* < Δ. Thus, taking Δ sufficiently small gives

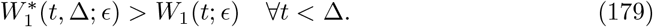

Moreover, note that, assuming Δ < *ϵ*^*′*^, if *t* > Δ is chosen such that *W*_1_(*t*; 1) < *ϵ*^*′*^ < *ϵ*, then

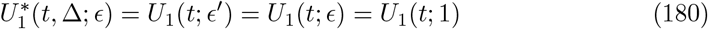

and hence

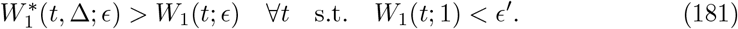

Finally, note that if *W*_1_(*t*; 1) ≥ *ϵ*^*′*^ then *W*_1_(*t*; *ϵ*^*′*^) = *ϵ*^*′*^ and hence

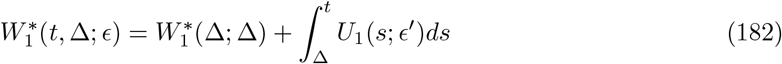

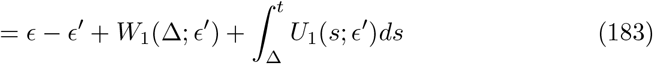

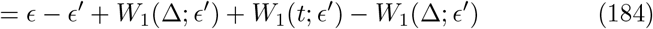

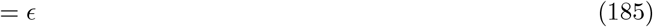

and so

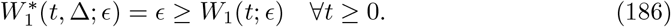

Moreover,

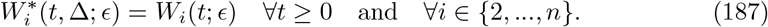

Thus, in particular, by Theorem 1, proved in [3], for each *i* ∈ {1, …, *n*},

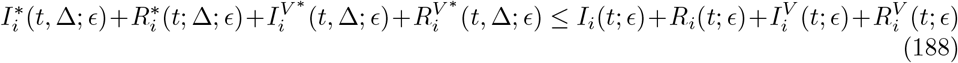

and

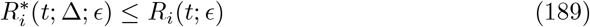

where the 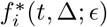 are the values of the model variables under the ***U*** ^*^(*t*, Δ; *ϵ*) vaccination policy and the *f*_*i*_(*t*; *ϵ*) are their values under the ***U*** (*t*; *ϵ*) vaccination policy.

Now, for all Δ > 0 and all *f* and *i*

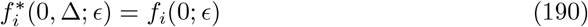

so, as all model variables except *S*_*i*_ and 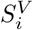 have derivatives that are bounded independently, there exists some *L* such that, for all

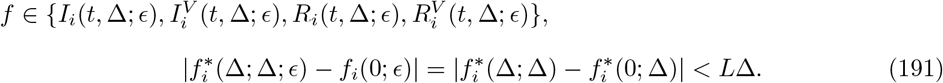

Moroever, thesame for *f* (0; *ϵ*) and *f* (0; *ϵ*^*′*^) except in the case of *S*_1_(0; *ϵ*). Thus,

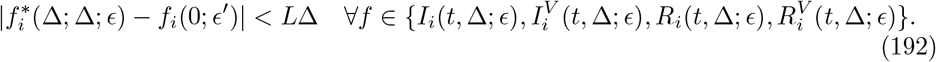

As only the *W*_1_ policy has an unbounded derivative in the Δ → 0 limit, it is also true that

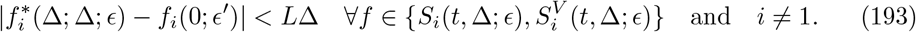

Moreover, note that (here suppressing the dependence on *ϵ*)

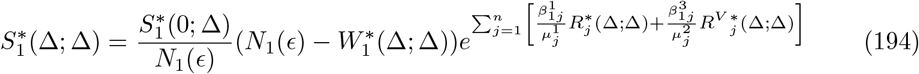

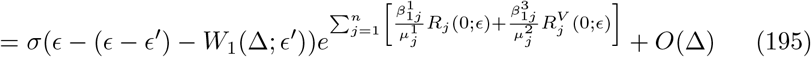

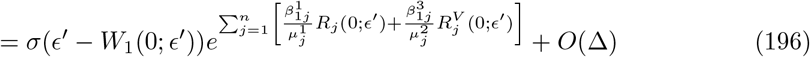

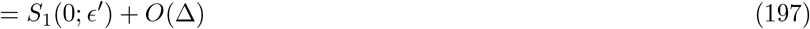

and hence,

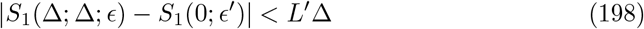

for some *L*^*′*^ > 0. Now, as (again suppressing the dependence on *ϵ*)

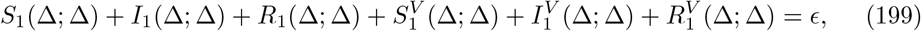

it is necessary that

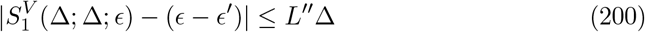

for some *L*^*′′*^ > 0. Thus, in particular, the values of the model variables 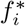 at time Δ converge to the initial conditions of the *ϵ*^*′*^ case, except that

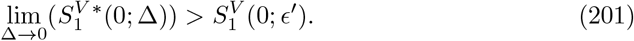

Moreover, note that for any *t* ≥ 0,

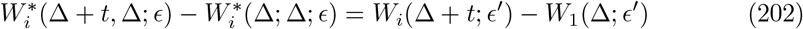

and so, as ***U*** ^*^ is bounded in [Δ, ∞)

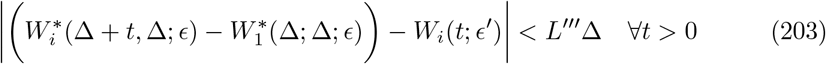

for some *L*^*′′′*^. Thus, define variables with a hat to denote those from the disease trajectory with initial conditions given by

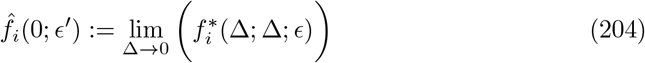

and with vaccination policy given by *W*_*i*_(*t*; *ϵ*^*′*^). Then, by considering the starred variables to come from an epidemic started at time *t* = Δ, Lemma 4.7 shows that

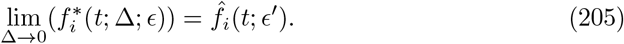

Thus, one can take the Δ → 0 limit in (188) and (189) to show

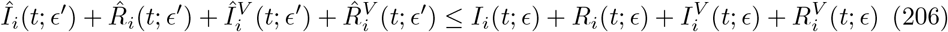

and

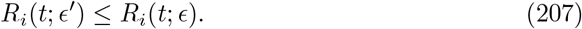

Taking *t* → ∞ in these inequalities shows that

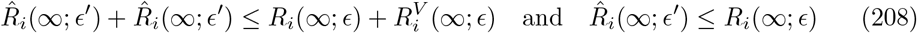

and hence, for any *κ*_*i*_ ∈ [0, 1],

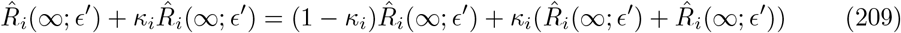

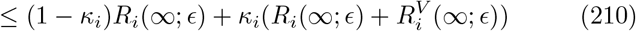

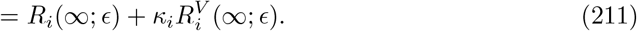

Summing these inequalities over *i* gives

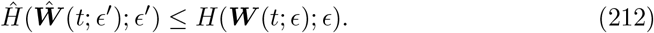

Finally, note that by Lemma 4.9, as the only change between cases *Ĥ* and *H* is an increase in one of the values of *S*^*V*^,

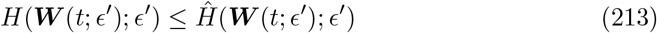

which, combined with (212) completes the proof of this proposition.

##### Theorem 2

This allows the overall proof of Theorem 2. The proof will rely on Theorem 1, which allows the creation of an *O*(1) decrease in the objective function by reducing *ϵ*. By comparing a sequence of policies satisfying (2) with a sequence that does not satisfy (2) and using Proposition 2.1, one can then create a sequence of optimal policies such that the associated sequence of objective values decreases by at least a fixed quantity at each step (and thus will eventually become negative, giving a contradiction).

Suppose (for a contradiction) that Theorem 2 does not hold for some fixed *α, τ* and *w*. Thus, for any *η* > 0, there is an *ϵ* ∈ (0, *η*) such that, for some ***U*** satisfying (2),

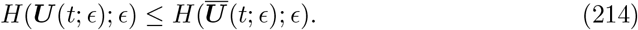

By optimality of ***Ū***(*t*; *ϵ*), (214) must in fact be an equality, and so it can be assumed that ***U*** (*t*; *ϵ*) = ***Ū***(*t*; *ϵ*), which will be done in the remainder of this proof (that is, if for some *ϵ* there is an optimal solution satisfying (2), then it will be assumed that ***Ū*** satisfies (2)). Thus, there is some *ϵ*_0_ such that

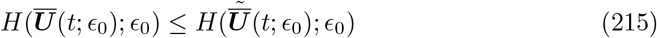

where 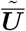 is defined by (4). Now, for *ϵ* < *ϵ*_0_, define ***U*** ^0^(*t*; *ϵ*) by

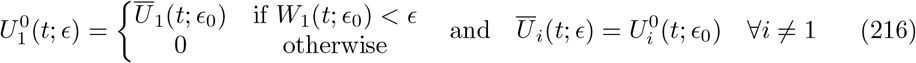

and note that this means that

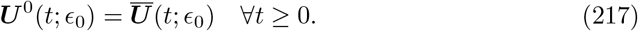

By (163) in the proof of Theorem 1, there exists some *δ*_1_ > 0 such that, for all *ϵ* < *δ*_1_,

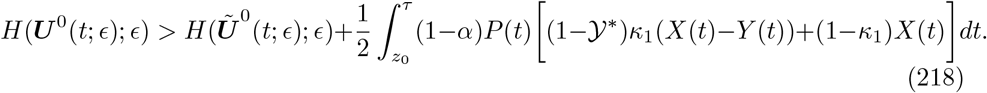

where

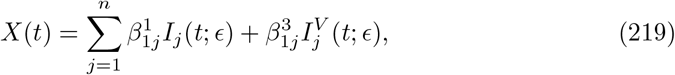

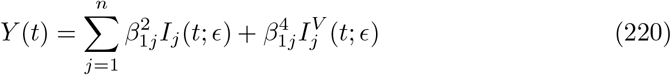

and

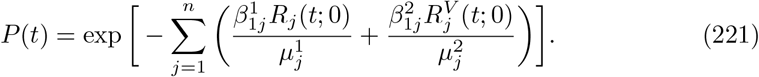

Note that *ρ*_0_, *τ* and 𝒴^*^ are independent of ***U*** ^0^, but *X*(*t*), *Y* (*t*) and *P* (*t*) are not. However, note that

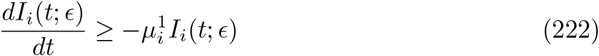

and so

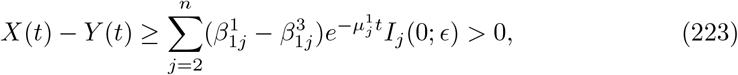

by the assumption (170), giving a bound that is independent of ***U*** ^0^. Moreover,

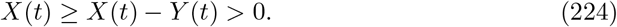

Finally, for *ϵ* ≤ 1,

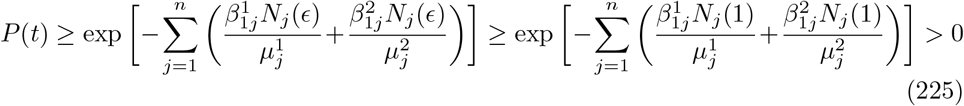

and this bound is again independent of ***U*** ^0^. Thus,

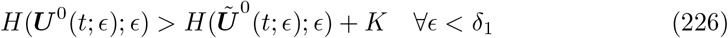

for some constant *K* > 0 where this is now independent of ***U*** ^0^. Now, by assumption, there must exist some *ϵ*_1_ ∈ (0, *δ*_1_) such that ***Ū***(*t*; *ϵ*_1_) meets the conditions (2) so

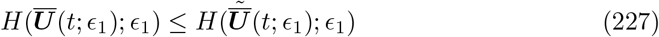

while by optimality

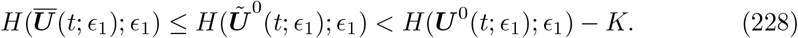

Now, moreover, note that by Proposition 2.1,

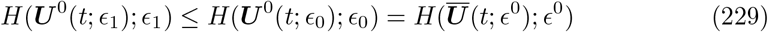

and so

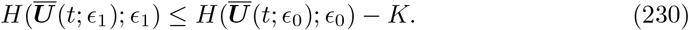

Now, this can be continued iteratively so that, for any *n* ≥ 0,

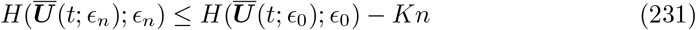

However, this means that eventually, one finds

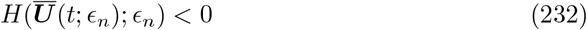

which is a contradiction. Thus, for each fixed *α, w* and *τ*, there must exist some *η* such that for any *ϵ* ∈ (0, *η*), the optimal solution does not satisfy (2).

Now, suppose that 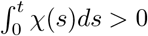 and suppose ***Ū***(*t*; *ϵ*) is an optimal solution for each value of *ϵ* such that, for each *t*

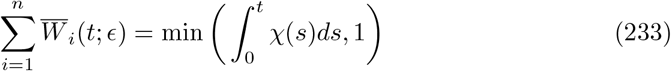

(note that this can be assumed by Theorem 2 in [3]). Now, suppose that, for some *t*

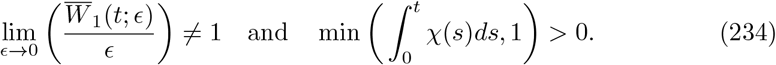

This means that there exists some *δ* > 0 such that there is a subsequence *ϵ*_*m*_ satisfying

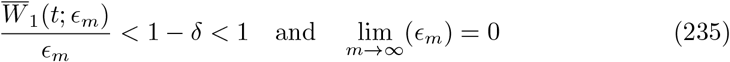

noting that

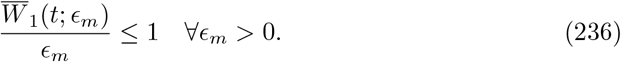

However, this means that for each *m*, ***Ū***(*t*; *ϵ*_*m*_) satisfies the condition (2) with *τ* = *t, α* = 1 − *δ* and 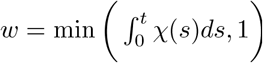. This is a contradiction to the previous part of the proof (as lim_*m*→∞_(*ϵ*_*m*_) = 0) and hence

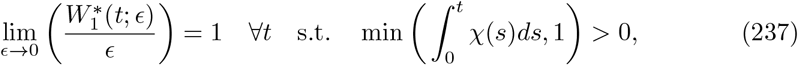

as required.

### 3 Proof of Theorem 3

Recall the definitions from the main text.

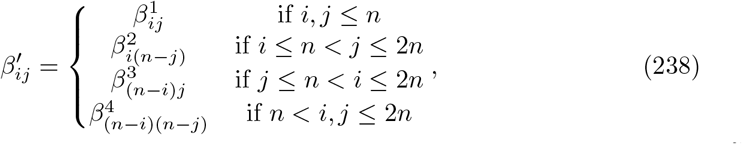

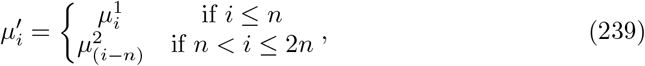

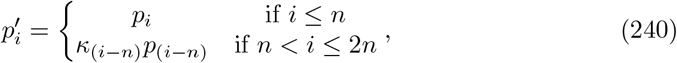

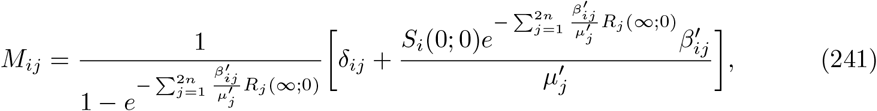

and

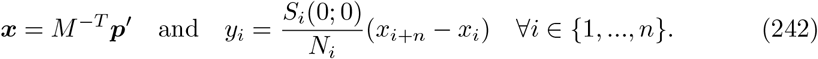

#### Theorem 3

*Suppose that, for all ϵ* > 0

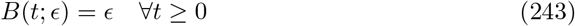

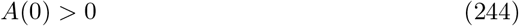

*and that the matrix M is invertible. Assuming that ϵ is sufficiently small so that it exists, define*

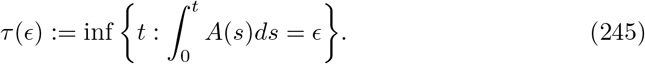

*Suppose that* ***U*** *satisfies the condition*

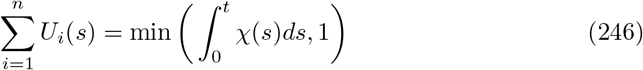

*where χ is defined in (169). Then, for sufficiently small ϵ, the objective function is given by*

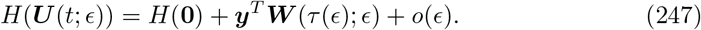

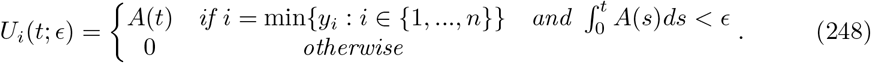

#### Proposition 3.1

Note that the *n*-group model can be considered as a 2*n*-group model once vaccination has finished - an idea that is formalised in the below proposition.

##### Proposition 3.1

*Define for i* ∈ {1, …, *n*},

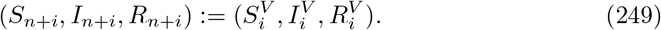

*Define further*

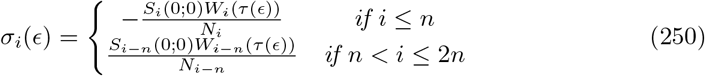

*and*

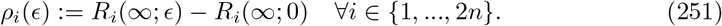

*Then, ρ*_*i*_(*ϵ*) *is o*(1) *as ϵ* → 0 *and*

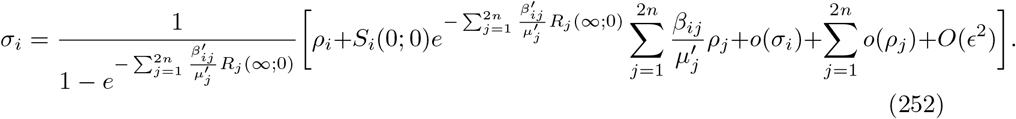

**Proof:** As *A* is continuous, there is some region (0, *δ*) such that

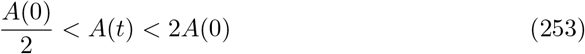

and hence

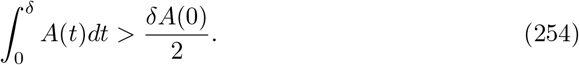

This lower bound is independent of *ϵ* and hence, for sufficiently small *ϵ*,

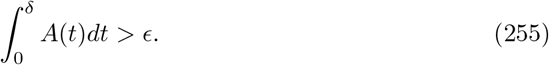

Now, by assumption,

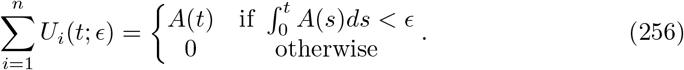

By continuity and the definition of *τ* (*ϵ*),

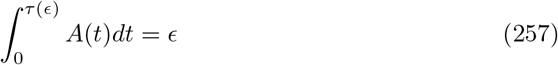

and note that it is necessary that *τ* (*ϵ*) = *O*(*ϵ*) as

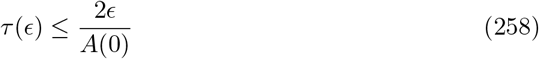

for sufficiently small *ϵ*.

Now, all of the variables are bounded independently of *ϵ* in the interval [0, *τ* (*ϵ*)] (including ***U***, which is bounded by 2*A*(0)). Moreover, assuming *N*_*i*_ > 0 for each *i* ∈ {1, …, *n*},

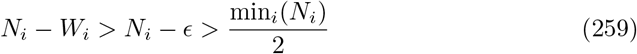

for sufficiently small *ϵ*. Thus, in particular, all of the derivatives of the model variables are bounded and so

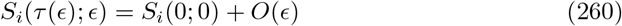

with analogous results for the other model variables, noting that the initial conditions are identical in each case. Thus, in particular,

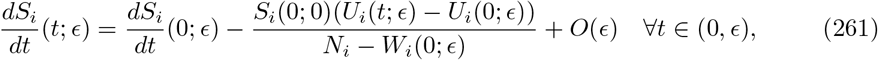

noting that the *U*_*i*_(*t*; *ϵ*) are the only quantities that can change by an *O*(1) amount in *O*(*ϵ*) time. Now, one can set *U*_*i*_(0; *ϵ*) = 0 to reduce notation (noting that the model depends only on the integral of *U*_*i*_). Moreover, as *W*_*i*_(0; *ϵ*) = 0, the initial conditions are independent of *ϵ* and *τ* (*ϵ*) = *O*(*ϵ*), integrating gives

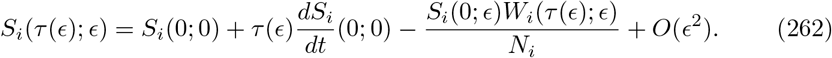

Similarly,

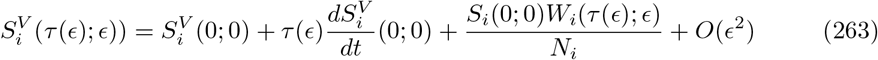

while for the other model variables, *f*_*i*_, there is no *O*(1) change to the derivative so

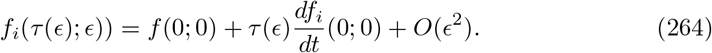

Now, for times *t* ≥ *τ* (*ϵ*), one has *U*_*i*_(*t*; *ϵ*) = 0 and so a standard multi-group SIR model (with initial conditions given by the model variables evaluated at time *τ* (*ϵ*)) is recovered. Thus in particular, the final number infected can be formulated in terms of a final size equation as follows. Define, for *i* ∈ {1, …, *n*},

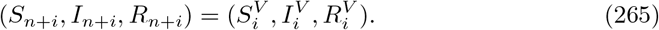

This new 2*n* group model has the same behaviour as the original model if the parameters are

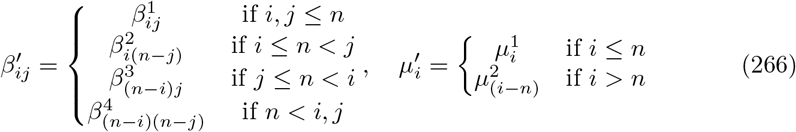

and

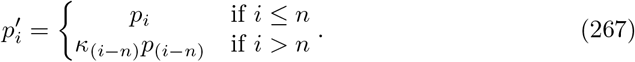

Thus, integrating the *S*_*i*_ equation between *τ* (*ϵ*) and *t* + *τ* (*ϵ*) gives

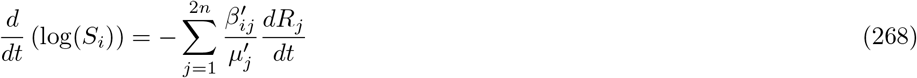

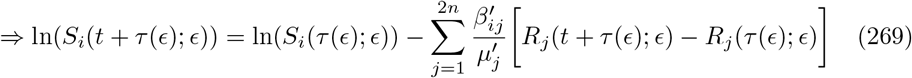

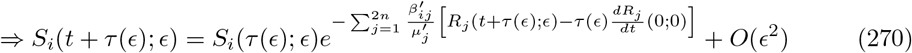

as *R*_*j*_(0; 0) = 0 for each *j*. Now, note that for any *t* ≥ 0,

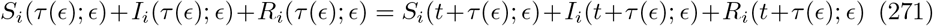

and hence, taking *t* → ∞ and using Lemma 4.3 shows that

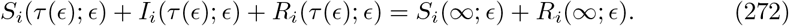

Hence, by (264),

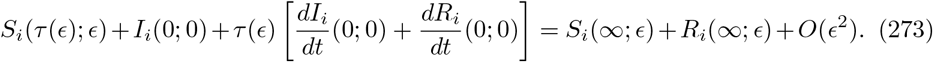

Now, substituting this into the limit of (270) as *t* → ∞ shows that

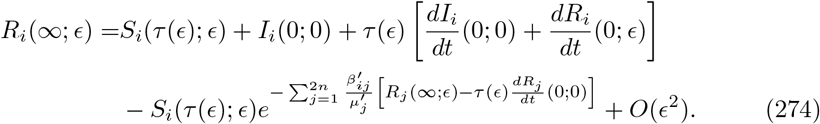

By treating this model as a model that has initial conditions given by the variable values at time *τ* (*ϵ*), one sees that these initial conditions differ from the initial conditions of the *ϵ* = 0 model by *O*(*ϵ*) (where no vaccination occurs in either case). This means that Proposition 1.2 can be used (as the vaccination policies ***U*** must have uniformly bounded finite support for sufficiently small *ϵ*) and so there exists some function *δ*(*ϵ*) such that, for all sufficiently small *ϵ*,

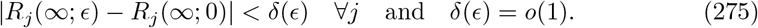

Thus, in particular, one can define functions *ρ*_*j*_(*ϵ*) such that

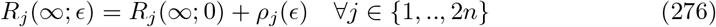

and

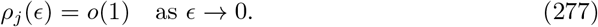

Furthermore, defining *σ*_*i*_ such that

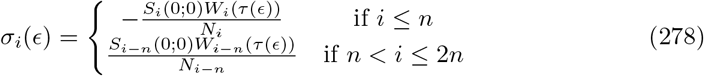

gives

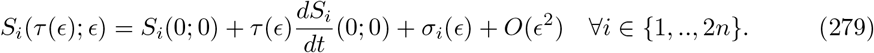

Now, when *σ*_*i*_(*ϵ*) = 0 for all *i*, it must be the case that *ρ*_*i*_(*ϵ*) = 0 for all *i* as the final size is unchanged (as no vaccination has taken place). Thus, in this case, (274) can be linearised to give

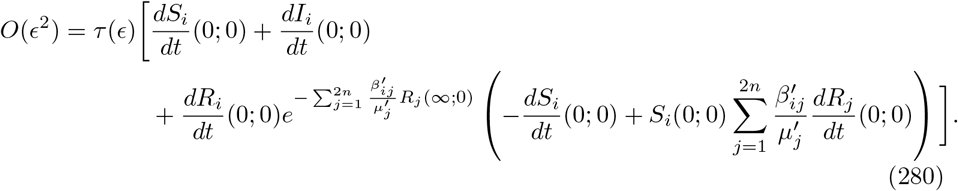

Note that this equality does indeed hold, as in the no vaccination case

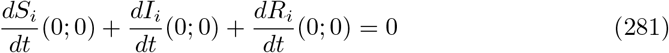

is the conservation of population law, while

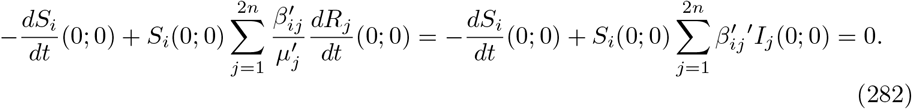

This means that, for non-zero *σ*_*i*_, all terms not dependent on *σ*_*i*_ or *ρ*_*i*_ cancel and so the linearisation becomes

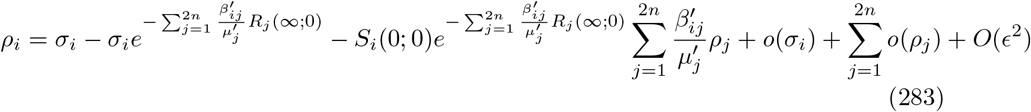

and so

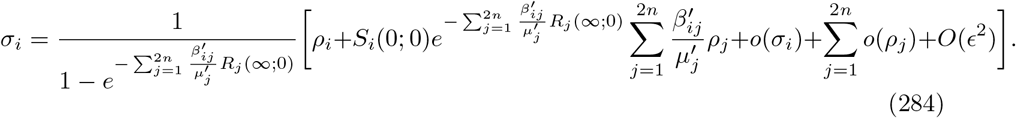

as required.

#### Proposition 3.2

The result of Proposition 3.1 can be written as a system of equations for vectors ***σ*** and ***ρ***

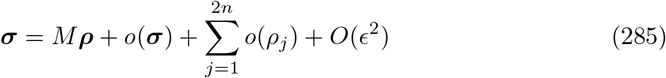

for some matrix *M* with non-zero determinant by assumption. However, it is important to establish the dominant balance in these equations, which is done through the following proposition

##### Proposition 3.2

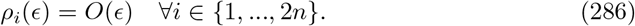

**Proof:** Suppose that this does not hold. Thus, there must be some sequence *ϵ*_*m*_ such that, for some *i*

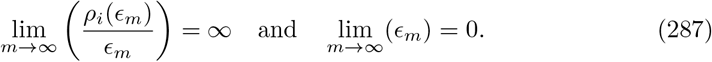

Define *J*^*^(*ϵ*) such that

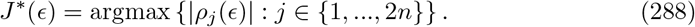

Now, by the finiteness of {1, …, 2*n*}, there exists some subsequence 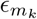 and some fixed *J* ∈ {1, …, 2*n*} such that

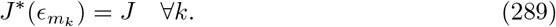

For notational convenience, assume that the original sequence *ϵ*_*m*_ has this property. Note that

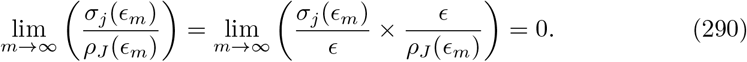

as *σ*_*j*_(*ϵ*) = *O*(*ϵ*) and *ϵ* = *o*(*ρ*_*i*_(*ϵ*)) ≤ *o*(*ρ*_*J*_ (*ϵ*)). Moreover,

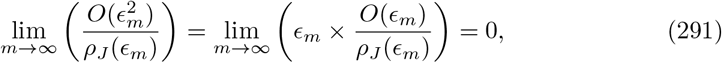

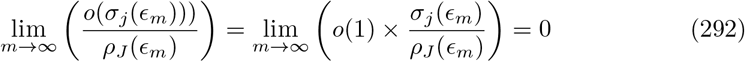

and

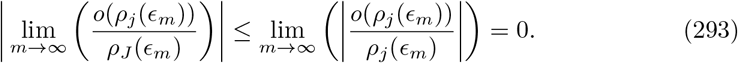

Note that there is some abuse of notation in these calculations, but, for example, an *O*(*ϵ*^2^) term in the limit represents any function which is *O*(*ϵ*^2^). Thus, dividing (285) by *ρ*_*J*_ (*ϵ*_*m*_) and taking *m* to ∞ shows that

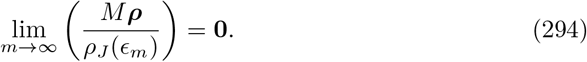

Define

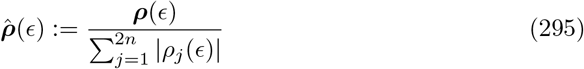

and note that

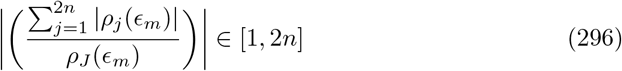

and thus remains finite and non-zero. Thus

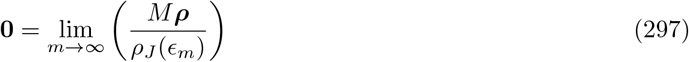

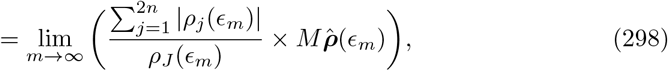

which means

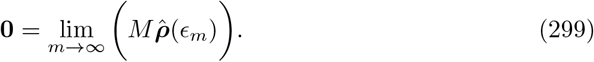

However, note that

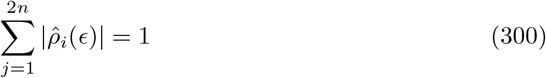

and hence the sequence 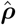 is bounded. Thus, by the Bolzano-Weierstrass Theorem, there must be some subsequence *m*_*k*_ such that 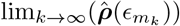 exists and is equal to some ***ρ***^*^ where

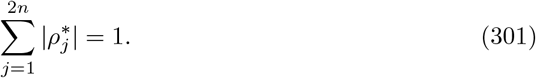

However, then, by continuity and the fact that *M* is invertible,

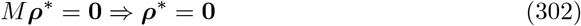

which is a contradiction to (301) as required. Thus, it must be the case that *ρ*(*ϵ*) = *O*(*ϵ*)

##### Theorem 3

Combining Proposition 3.2 with the fact that *σ*_*i*_ = *O*(*ϵ*) means that (285) can be written as

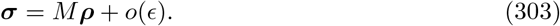

Thus, one can multiply the equation by *M*^−1^ to get

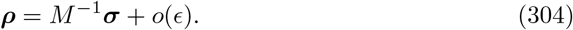

Hence, given vectors ***p*** and ***q*** where

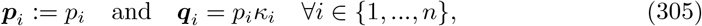

the change to the objective function is given by

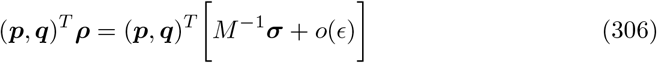

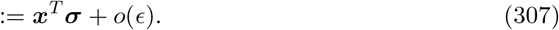

Now, note that, for *i* ∈ {1, …, *n*},

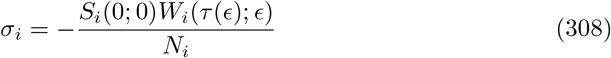

while, for *i* ∈ {*n* + 1, …, 2*n*}

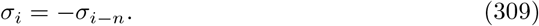

Hence, one can write (307) as

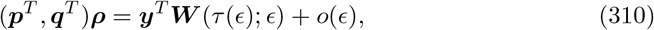

where

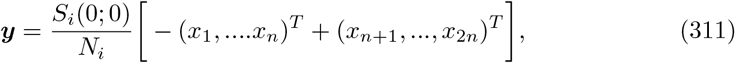

as required by Theorem 3. The only restriction is that all the *W*_*i*_ are non-negative and that

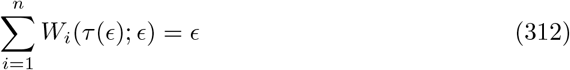

and so the optimisation problem becomes

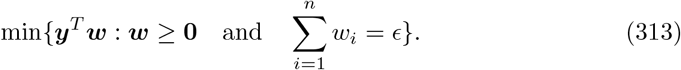

Now, by Theorem 1, proved in [3] and stated in the appendices, it must be the case that the objective function is non-increasing in ***w***. Thus, in particular, one must have

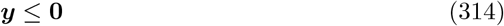

as otherwise, if *y*_*i*_ > 0 then setting ***w*** = *ϵ****e***_*i*_ (where ***e***_*i*_ is the ith canonical basis vector) means that

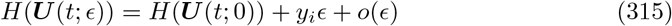

and so, for sufficiently small *ϵ*,

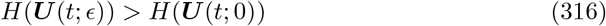

which is a contradiction. Hence, ***y*** ≤ **0** which means that the optimisation problem is an example of a continuous knapsack problem and one can readily see that a solution given is by

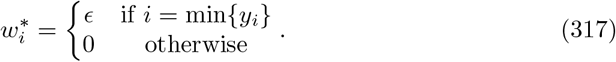

As this minimum is unique by assumption, this is the unique leading order optimal solution to the optimisation problem.

A technical note is that this only proves the form of the optimal solution to leading order. Indeed, if

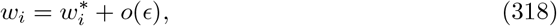

then the optimal objective value is unchanged to leading order. Hence, this restriction is given in the statement of the theorem (although in practice is unimportant).

### 4 Supplementary lemmas

This section contains the supplementary lemmas that have been used in the proofs of Theorems 1-3. All but two of these lemmas were proved in [3] and so their proofs will not be reproduced here, but they have been included for completeness and for ease of access. The exceptions are Lemma 4.8 and 4.9.

#### Lemma 4.1

##### Lemma 4.1

*Consider a continuous, time-dependent, matrix A*(*t*) *which satisfies*

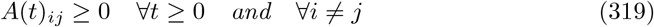

*and a constant matrix B that satisfies*

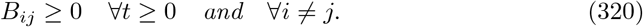

*Then, suppose that each element of A*(*t*) *is non-increasing with t and that*

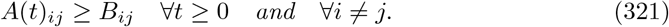

*Moreover, define an initial condition* ***v*** *and suppose that* ***y*** *and* ***z*** *solve the systems*

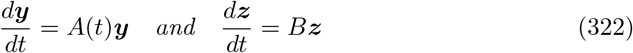

*with*

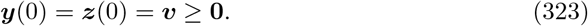

*Then*,

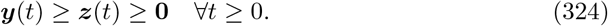

**Proof:** This was proved as Lemma B.2 in [3]

#### Lemma 4.2

##### Lemma 4.2

*Define the set of functions*

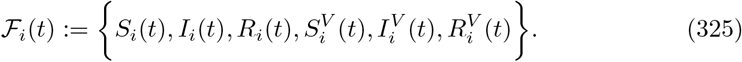

*Then, for all t* ≥ 0 *and i* ∈ {1, …, *n*},

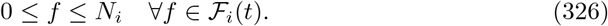

**Proof:** This was proved as Lemma B.3 in [3].

#### Lemma 4.3

##### Lemma 4.3

*For each i*,

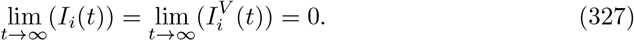

**Proof:** This was proved as Lemma B.4 in [3].

#### Lemma 4.4

##### Lemma 4.4

*Suppose that I*_*i*_(*t*) > 0 *for some t* ≥ 0 *and some i* ∈ {1, …, *n*}. *Then*

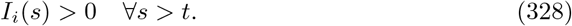

*An analogous result holds for* 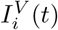.

**Proof:** This was proved as Lemma B.5 in [3].

#### Lemma 4.5

##### Lemma 4.5

*Define*

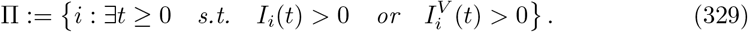

*Moreover, define*

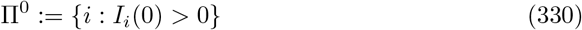

*and the n by n matrix M by*

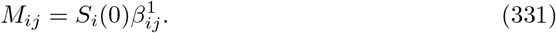

*Then, define the connected component C of* Π^0^ *in M as follows. The index i* ∈ {1, …, *n*} *belongs to C if any only if there is some sequence a*_1_, …, *a*_*k*_ *such that*

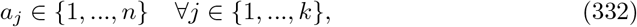

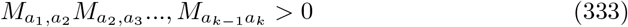

*and*

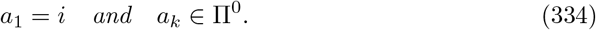

*Then*,

a. *i* ∈ *C* ⇒ *I*_*i*_(*t*) > 0 *t* > 0. ∀*t* > 0.
b. Π = *C* ⋃ Π^0^.

*Thus, in particular*,

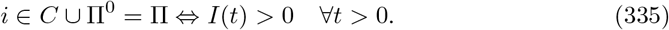

**Proof:** This was proved as Lemma B.6 in [3].

#### Lemma 4.6

##### Lemma 4.6

*Suppose that f* : *ℜ*^*n*^ → *ℜ is differentiable with bounded derivatives. Define C to be any closed bounded subset of ℜ*^*n*^. *Then, f is Lipschitz continuous on C - that is, there exists some L* > 0 *such that*

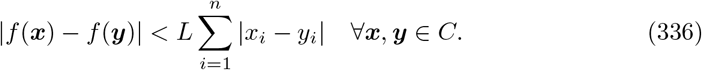

**Proof:** This was proved as Lemma B.7 in [3].

#### Lemma 4.7

##### Lemma 4.7

*Define the set of functions*

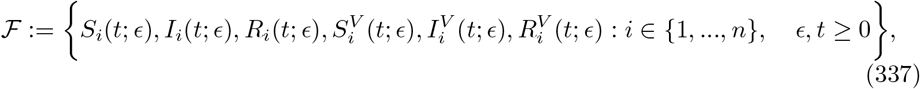

*where for each fixed ϵ, these functions solve the model equations with parameters*

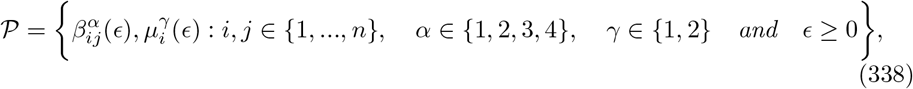

*initial conditions*

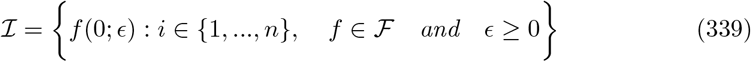

*and vaccination policy* ***U*** (*t*; *ϵ*). *Suppose further that the population sizes are independent of ϵ, except in group 1 where N*_1_(*ϵ*) *satisfies*

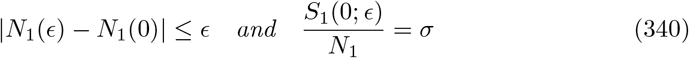

*for some constant σ*.

*Suppose that*

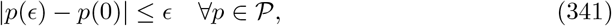

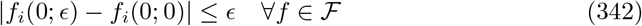

*and that*

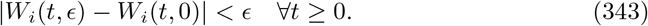

*Moreover, suppose that for each i* ∈ {1, …, *n*} *and ϵ* ≥ 0,

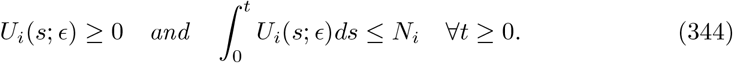

*Then, for each δ* > 0 *and each T* > 0 *there exists some η* > 0 *(that may depend on T and δ) such that*

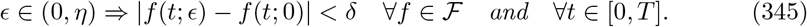

**Proof:** An almost identical result was proved in Lemma B.8 from [3], with the only exception being that *N*_1_ can vary in this example. However, note that by replacing 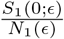 with *σ*, this lemma can be identically.

#### Lemma 4.8

##### Lemma 4.8

*Suppose that i* ∈ Π, *with* Π *defined as in Lemma 4*.*5. Then*,

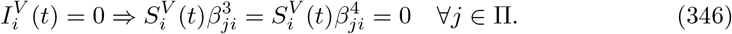

**Proof:** Suppose that there exists some *t* and some *i, j* ∈ Π such that

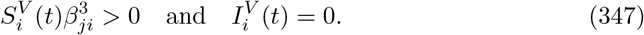

Then, by continuity, there exists some *a* < *t* such that

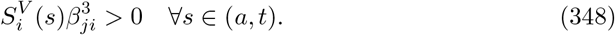

Moreover, by Lemma 4.4, it is necessary that

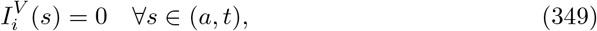

while, by Lemma 4.5

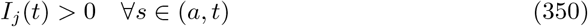

and hence (using the fact that 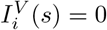 ∀*s* ∈ (*a, t*))

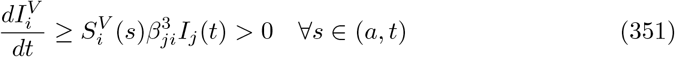

and so

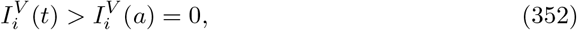

which is a contradiction as required. The final equality then follows as 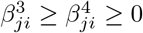.

#### Lemma 4.9

##### Lemma 4.9

*Suppose that the disease trajectories* ***S*** *and* 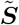 *are given by the same model equations, parameters, vaccination policy* ***U*** *and initial conditions except for the fact that*

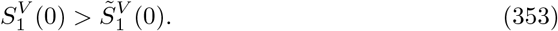

*Then, if the objective functions are denoted by H and* 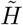 *for the two policies*

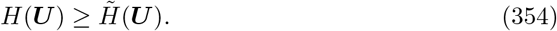

**Proof:** Define a new disease model, denoted by hats where a new group (*n* + 1) is added in such that its unvaccinated compartments behave like the vaccinated compartments of group 1 and its vaccinated compartments are perfectly immune from the disease. That is,

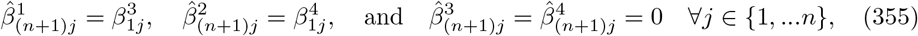

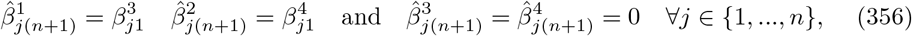

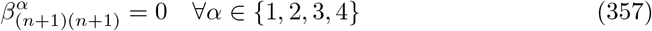

and

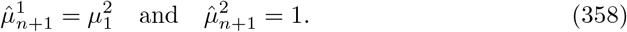

Suppose further that all other parameter values are identical, and that the only differences in the initial conditions is that

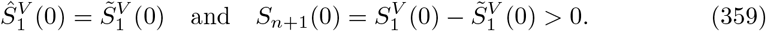

Then, note that

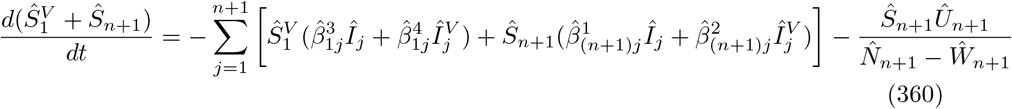

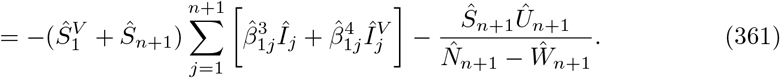

Moreover, for *i* ≠ 1

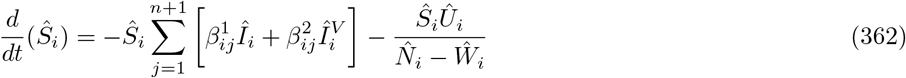

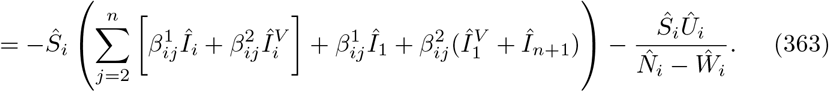

Thus, with similar calculations for 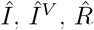 and 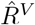, by the initial conditions and by the uniqueness of solution, in the case that *Û*_*n*+1_ = 0,.

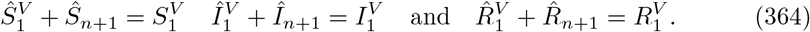

Thus, setting

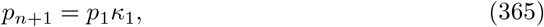

this means that

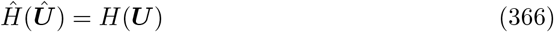

for any ***Û*** such that *Û*_*n*+1_ = 0 and *Û*_*i*_ = *U*_*i*_ for any *i* ≠ *n*.

Now, define a vaccination policy ***Û*** *(*t*; Δ) such that

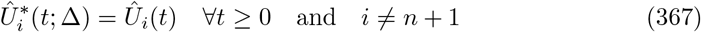

and

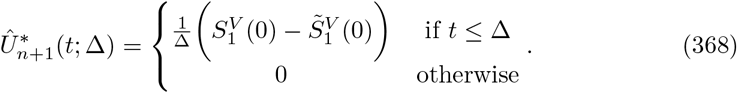

Then, this means that

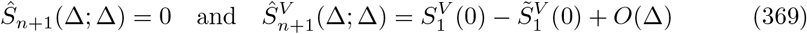

while all other variable values at time Δ differ by at most *O*(Δ) from their initial values. Thus, define by an overbar the model given by the initial conditions which are the same as those in the hat model, but with

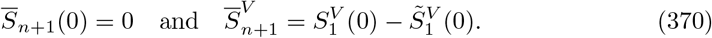

Suppose also that the vaccination policy in this case is equal to ***U***, which is the pointwise limit of the vaccination policy ***Û*** *(*t*; Δ) (for *t* > 0). Then, using Proposition 1.2, by considering the values of the variables 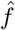 at time Δ to be the initial conditions, one finds that for any finite time *t*,

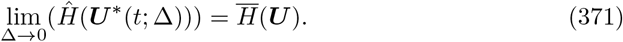

Note this holds as it is assumed that ***U*** is bounded and so

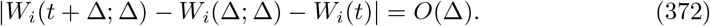

Moreover, note that the only difference between the bar model and the tilde model is in group (*n* + 1). However, by the fact that 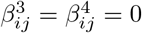 if (*n* + 1) ∈ {*i, j*}, the value of 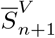 is constant and the other variable values are independent of it. Thus, by the uniqueness of solution, this means that

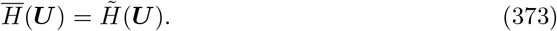

Finally, note that by Theorem 1, it must be necessary that for any Δ > 0

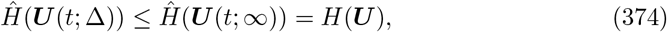

where Δ = ∞ corresponds to no vaccination taking place in group (*n* + 1) (and hence the original objective function *H* is recovered). Thus,

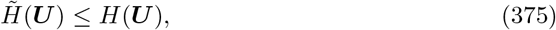

as required.

#### Theorem 1

##### Theorem 1

*Suppose that* ***U*** *and* ***Ũ*** *are feasible, bounded, Lebesgue integrable vaccination policies. Suppose further that for each i* ∈ {1, …, *n*} *and t* ≥ 0

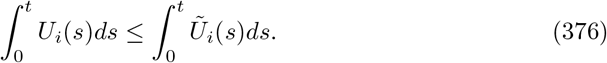

*Then, for each t* ≥ 0 *and i* ∈ {1, …, *n*}

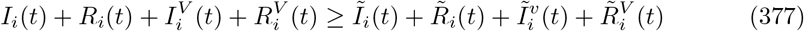

*and*

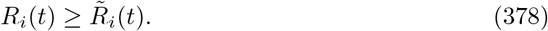

*Moreover*,

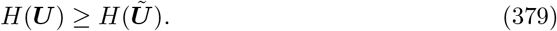

**Proof:** A proof of this theorem is given in [3], where it is Theorem 1. Note that the first two results are not in the statement of Theorem 1 in [3], but can be found at the end of the proof.

### Theorem 2

#### Theorem 2

*Suppose that B is differentiable, and that there is an optimal solution* ***U***. *Then, define the function*

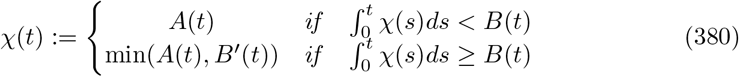

*and suppose that χ*(*t*) *exists and is bounded. Then, there exists an optimal solution* ***Ũ*** *such that*

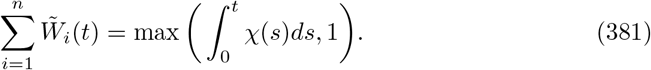

*Moreover, if χ*(*t*) *is continuous almost everywhere, there exists an optimal solution* ***Ũ*** *such that*

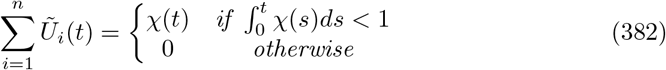

**Proof:** A proof of this theorem is given in [3] where it is Theorem 2.

